# Classification of virologic trajectories during nucleos/tide analogue treatment of hepatitis B virus (HBV) infection

**DOI:** 10.1101/2023.12.01.23299288

**Authors:** Tingyan Wang, Cori Campbell, Alexander J Stockdale, Stacy Todd, Karl McIntyre, Andrew Frankland, Jakub Jaworski, Ben Glampson, Dimitri Papadimitriou, Luca Mercuri, Christopher R Jones, Hizni Salih, Gail Roadknight, Stephanie Little, Theresa Noble, Kinga A Várnai, Cai Davis, Ashley I Heinson, Michael George, Florina Borca, Louise English, Luis Romão, David Ramlakhan, NIHR HIC Viral Hepatitis and Liver Disease Consortium, Kerrie Woods, Jim Davies, Eleni Nastouli, Salim I Khakoo, William Gelson, Graham S Cooke, Eleanor Barnes, Philippa C Matthews

## Abstract

**Background & Aims:** The dynamics of HBV viral load (VL) in patients with chronic hepatitis B (CHB) on nucleos/tide analogue (NA) treatment and its relationship with liver disease are poorly understood. We aimed to study longitudinal VL patterns and their associations with CHB clinical outcomes.

**Methods:** Utilising large scale, routinely collected electronic health records from six centres in England, collated by the National Institute for Health and Care Research Health Informatics Collaborative (NIHR HIC), we applied latent class mixed models to investigate VL trajectory patterns in adults receiving NA treatment. We assessed associations of VL trajectory with alanine transaminase (ALT), and with liver fibrosis/cirrhosis.

**Results:** We retrieved data from 1885 adults on NA treatment (median follow-up 6.2 years, interquartile range (IQR) 3.7-9.3 years), with 21,691 VL measurements (median 10 per patient, IQR 5-17). Five VL classes were identified from the derivation cohort (n=1367, discrimination: 0.93, entropy: 0.90): class 1 ‘long term suppression’ (n=827, 60.5%), class 2 ‘timely virological suppression’ (n=254, 18.6%), class 3 ‘persistent moderate viraemia’ (n=140, 10.2%), class 4 ‘persistent high-level viraemia’ (n=44, 3.2%), and class 5 ‘slow virological suppression’ (n=102, 7.5%). The model demonstrated a discrimination of 0.93 and entropy of 0.88 for the validation cohort (n=518). ALT decreased variably over time in VL-suppressed groups (classes 1, 2, 5; all p<0.001), but did not significantly improve in those with persistent viraemia (classes 3, 4). Patients in class 5 had 2-fold increased hazards of fibrosis/cirrhosis compared to class 1 (adjusted hazard ratio, 2.00; 95% CI, 1.33-3.02).

**Conclusions:** Heterogeneity exists in virological response to NA therapy in CHB patients, with over 20% showing potentially suboptimal responses. Slow virological suppression is associated with liver disease progression.

## Introduction

Chronic hepatitis B (CHB) is a significant global health concern, affecting nearly 300 million individuals worldwide, and accounting for a large proportion of the global burden of cirrhosis and hepatocellular carcinoma (HCC) (1). In 2019, the World Health Organization (WHO) estimated there were approximately 1.5 million new infections and 820,000 deaths related to Hepatitis B Virus (HBV) worldwide (2,3). International sustainable development goals for the elimination of HBV as a public health threat by 2030 aim for 65% reduction in mortality and 90% reduction in incidence compared with baseline levels observed in 2015 (4). Successful deployment of oral antiviral treatment is a cornerstone of interventions required to achieve these ambitious targets.

Long-term antiviral treatment with nucleos/tide analogue (NA) agents suppresses HBV replication and reduces the long-term risk of inflammatory liver disease, cirrhosis, and HCC (5,6). However, complete virologic suppression can be slow (7–10), potentially taking up to 2-3 years for HBV DNA viral load (VL) to suppress below the limit of detection in blood. Furthermore, in a minority of individuals, there is a risk of persistent viraemia or viral rebound (7–12), which might be associated with high pre-treatment HBV DNA levels (7), HBeAg positivity at baseline (8,9), HIV coinfection (11), incomplete treatment adherence (12), and/or HBV resistance to antiviral therapy (13,14). While studies have investigated the overall pattern of changes in VL (e.g. time to virological response, the proportion of non-suppressed) during antiviral treatment (7–10), the detailed characterisation of longitudinal VL trajectory patterns has been limited, and largely based on clinical trial cohorts which may not represent real-world populations.

At present, only a minority of the population with CHB are deemed eligible for NA treatment, based on guidelines for HBV management which use clinical and laboratory assessments to identify those at highest risk (5,6,15,16). However, such guidance is changing over time, recognising the potential benefits of relaxed and simplified treatment criteria, as exemplified by new recommendations in China which suggest treatment for all those with detectable HBV DNA and ALT above the upper limit of normal (ULN) (17). As an increasing number of individuals embark on NA therapy, there is a pressing need to understand therapeutic responses, and to identify and interpret the clinical significance of situations in which virologic suppression is inadequate.

There are recommendations in European (16) and UK guidelines (6) for supporting management of patients in whom viraemia is persistent or rebounds on NA treatment. These suggest reviewing and supporting adherence in people taking tenofovir disoproxil fumarate (TDF) in whom virological suppression is not achieved (particularly in the setting of advanced liver disease) or in whom there is not an ongoing downward trajectory of VL over time. In such instances, adding lamivudine (3TC) or entecavir (ETV) to tenofovir can also be considered (16). However, there is a lack of evidence or structured algorithms to determine the optimal timing for assessing the impact of such an intervention (e.g. 48 vs. 96 weeks) and to establish a VL threshold associated with clinical response (e.g. <2000 IU/ml). Thus, more evidence is needed to identify patients at risk of inadequate virologic suppression, to inform investigations or interventions, and to assess the public health consequences of incomplete treatment response.

Latent class mixed modeling, a contemporary unsupervised approach, has already demonstrated clinical relevance in various disease areas, including cardiovascular disease (18), chronic kidney disease (19), human immunodeficiency virus (HIV) (20), and coronavirus disease (caused by SARS-CoV-2) (21), by disaggregating sub-phenotypes. We used this approach to avoid any prior assumptions about HBV VL trajectories and to take an unbiased approach to identify distinct on-treatment groups. We took advantage of the opportunity of large-scale clinical data collected from secondary care services in the UK through the National Institute for Health and Care Research (NIHR) Health Informatics Collaborative (HIC) programme.

Our objectives include a) characterising HBV virologic trajectory patterns in adults receiving NA therapy using large-scale electronic patient records (EPR), b) stratifying demographic, laboratory, and clinical characteristics by VL trajectory pattern, c) assessing liver inflammation across VL trajectory patterns, and d) determining whether identified VL patterns were associated with liver disease progression. We aimed to provide insights into the dynamics of HBV virologic trajectories during NA therapy, which are relevant to supporting the management of individuals on treatment, facilitating the development of personalised treatment approaches, and providing evidence that can inform public health approaches.

## Patients and methods

### Data collection

We used routinely collected clinical data from 10,460 individuals under longitudinal follow up for HBV infection in secondary care services. These were collated from six National Health Service (NHS) trusts (distinct regional organisations, each a separate legal entity, responsible for provision and commissioning of health care, and each made up of several hospitals) in England, established through the NIHR HIC Viral Hepatitis and Liver Disease theme: Cambridge University Hospitals NHS Foundation Trust (CUH), Imperial College Healthcare NHS Trust (ICHT), Liverpool University Hospitals NHS Foundation Trust (LUH), Oxford University Hospitals NHS Foundation Trust (OUH), University College London Hospitals NHS Foundation Trust (UCLH), and University Hospital Southampton NHS Foundation Trust (UHS). Data were collated by a local NIHR HIC team in each site, being drawn from operational systems including EPR systems into a data warehouse and linked to produce a comprehensive record for each patient with a data validation process, as described in a previous methods paper and cohort profile (22,23). Data were shared with the host organisation (Oxford University Hospitals NHS Foundation Trust), validated against the defined data model and then integrated into the central database for research studies. The management of the central database is governed by the NIHR HIC Data Sharing Framework.

The data elements used for this study included demographics, deprivation scores (Index of Multiple Deprivation (IMD) (24)), laboratory tests (liver biochemistry, other tests reflecting liver health, identification of other chronic viral infections) (**Table S1**), HBV treatment regimen information, imaging, liver biopsy reports and ICD diagnosis codes for inferring liver fibrosis and cirrhosis status.

### Eligibility criteria

To be included in this study, all of the following criteria had to be met: (i) HBV infection (positive HBsAg or detectable HBV DNA test), (ii) any record of antiviral treatment with NA (monotherapy or combination) therapy, including TDF or tenofovir alafenamide (TAF), ETV, and older regimens, e.g. lamivudine, adefovir, (iii) ≥2 quantified HBV DNA VL measurements spanning ≥6 months during the period of treatment, and (iv) aged ≥18 years at the first quantified HBV DNA VL.

### Statistical analysis

We aimed to use 70% of our sample for model development, and 30% for validation, and thus based on the number of patients for each site, data from four sites (ICH, LUH, UCLH, UHS) were collectively utilised as the derivation set ensuring a sufficient sample size for modelling, while data from the remaining two sites (CUH, OUH) were exclusively reserved for external validation. We defined the date of first VL measurement during the period of treatment as ‘baseline’, as earliest treatment start dates were not reliably recorded in the EPR systems. All analyses for this study were conducted in R 4.2.1. We conducted latent class mixed modelling (25) to characterise VL trajectories using the *lcmm* package (version 2.0.2).

In the study cohort, 150 individuals had VL reported as ‘detectable’ without a quantitative result at some point during their clinical record. This typically suggests a level of viraemia that is below the limit of quantification but at which the assay can still detect the presence of HBV DNA. However, in the majority of these (103/150) the unquantified value occurred in the middle of a longitudinal record of quantitative VL measurements and was therefore not likely to influence the class assigned by the model. To avoid assigning an arbitrary numerical value to qualitative results, we therefore excluded VL datapoints that had a qualitative-only result.

We applied a nonlinear (log) transformation to HBV DNA VL levels before modelling. Varied numbers of VL measurements at different time points were considered in the approach by specifying fixed-effects at the latent process level and class-specific level, as well as including random-effects for individual patients. We explored modelling with different functions of time (linear, quadratic, splines) and finally determined natural splines on time with internal knots (at first and second tertiles) and boundary knots (at 2.5th and 97.5th percentiles) as the most appropriate function of time. In the latent class mixed models, we also included age, sex, and ethnicity as covariates in the class-membership multinomial logistic model. Age was included by performing splines with an internal knot and boundary knots. For models with more than one class, we randomly generated the initial values from the asymptotic distribution of the estimates of the one-class model. We derived a set of models with a varying number of latent classes, and the optimal number of VL trajectory patterns (which we defined as ‘classes’) was determined by the Bayesian information criteria (BIC), the Akaike information criteria (AIC), the discrimination, the relative entropy, the odds of correct classification, and the interpretability of the model (19,26,27). The calculation of discrimination and entropy (28) is provided in **Table S2**.

We tabulated the demographic, laboratory, and treatment characteristics at baseline across the identified classes in derivation and validation cohorts. For categorical variables, chi-square or Fisher’s exact tests were used for comparison. For continuous variables, t or Wilcoxon tests were used for comparison. All significance tests performed were two-sided.

To determine whether persistent viraemia drives liver inflammation, we combined data from derivation and validation cohorts, and performed linear mixed effects modelling with random intercepts and slopes to assess longitudinal trends of alanine transferase (ALT) levels for these distinct patient classes. All linear mixed effects models were adjusted for demographics including age, sex, and ethnicity. We also provided distribution and stratification of ALT levels at baseline, and at 12, 24, 36, 48, 60, and 72 months after the baseline, in the five classes.

As described previously (23), we identified liver fibrosis and cirrhosis status based on the following data sources: Ishak or METAVIR scores from biopsy reports, liver stiffness measurements from transient elastography (FibroScan) if available, imaging reports (ultrasound, CT, and MRI), ICD diagnosis information, or on aspartate aminotransferase (AST) to platelet ratio index (APRI) or Fibrosis-4 (FIB-4) scores (which were calculated by laboratory tests using measurements of platelets, AST, and ALT measured within 12 weeks of each other). We used pre-defined thresholds for significant/advanced fibrosis and cirrhosis: 1.5 and 2.0 for APRI score, respectively; 3.25 and 3.6 for FIB-4 score, respectively (29,30). For the analysis of associations of VL trajectory classes with liver fibrosis and cirrhosis, we excluded individuals with no data available for liver fibrosis/cirrhosis diagnosis. To explore liver disease progression for patients with different VL trajectory classes, patients with fibrosis and cirrhosis occurring within 6-months of their first VL measurement (baseline) or follow up of liver disease status <6 months were excluded to avoid immortal time bias.

We compared the presence of liver fibrosis and/or cirrhosis in the overall cohorts (combining derivation and validation cohorts according to the VL trajectory classes). We performed Kaplan-Meier (K-M) analysis to compare the probability of being fibrosis and cirrhosis-free over time between classes. We used univariate and multivariate Cox proportional hazards models to investigate whether the distinct virologic classes predicted disease progression to liver fibrosis/cirrhosis, reporting 95% confidential intervals (CIs). To handle missingness on covariates, multiple imputations for missing data were performed using the MICE algorithm (31). We conducted Cox proportional-hazards modelling with the imputed datasets and pooled the estimates. We performed sensitivity analysis to investigate the robustness of associations observed. We undertook receiver operating characteristic (ROC) analysis using *pROC* package (32) to examine the predictive relationship between VL trajectory and liver fibrosis/cirrhosis, compared to that of other predictors of liver fibrosis and cirrhosis identified by multivariate analysis, reporting the area under the curve (AUC), sensitivity, specificity, and accuracy.

To explore the utility of the developed VL trajectory classification model for facilitating patient stratification in the real world, we conducted sensitivity analysis on the validation cohort with different scenarios, by changing the number of HBV DNA VL measurements per patient. We also assessed the performance of classification with and without restrictions on the minimum follow up duration of HBV DNA VL.

## Results

### Characteristics of study cohort

We included 1885 adults receiving NA treatment for chronic HBV infection, 62.8% male with median age 43 years (IQR 34-54). Longitudinal VL data were available over a median follow-up duration of 6.2 years (interquartile range [IQR], 3.7-9.3 years). We analysed a total of 21,691 VL measurements, with a median of 10 measurements per patient (IQR 5-17, range 2-45). The flow chart of participant selection is illustrated in **Figure S1**. Baseline characteristics and follow up information of the derivation and validation cohorts are presented in **Table 1**. Missingness of some laboratory parameters varied by sites (e.g. AST and HBeAg), reflecting differences in laboratory testing practice.

**Table 1.** Follow-up duration, demographics, and medical characteristics at presentation of adults with chronic HBV infection on NA therapy in the overall study cohort, and the derivation and validation cohorts.

| Parameter | Overall Study cohort | Derivation cohort | Validation cohort |
| --- | --- | --- | --- |
| Number of patients | 1885 | 1367 | 518 |
| Follow-up duration, years | 6.2 [3.7-9.3] | 6.6 [3.8-9.4] | 5.7 [3.5-7.4] |
| Total number of VL measurements | 21,691 | 16,980 | 4,711 |
| Number of VL measurements per patient | 10 [5-17] | 11 [6-18] | 8 [4-12] |
| Sex, male | 1184 (62.8) | 846 (61.9) | 338 (65.3) |
| Age, years | 43 [34, 54] | 44 [34, 55] | 41 [33, 52] |
| Age group, years |  |  |  |
| 18-24 | 77 (4.1) | 55 (4.0) | 22 (4.2) |
| 25-34 | 442 (23.4) | 304 (22.2) | 138 (26.6) |
| 35-44 | 498 (26.4) | 338 (24.7) | 160 (30.9) |
| 45-54 | 409 (21.7) | 315 (23.0) | 94 (18.1) |
| 55-64 | 284 (15.1) | 206 (15.1) | 78 (15.1) |
| 65-74 | 131 (6.9) | 113 (8.3) | 18 (3.5) |
| >=75 | 44 (2.3) | 36 (2.6) | 8 (1.5) |
| Ethnic group |  |  |  |
| Asian | 624 (39.4) | 418 (36.7) | 206 (46.3) |
| Black | 314 (19.8) | 243 (21.4) | 71 (16.0) |
| White | 393 (24.8) | 260 (22.8) | 133 (29.9) |
| Mixed/other ethnicity | 252 (15.9) | 217 (19.1) | 35 (7.9) |
| Not reported | 302 (16.0) | 229 (16.8) | 73 (14.1) |
| IMD (decile) | 5 [3, 7] | 4 [2, 6] | 6 [4, 9] |
| IMD category |  |  |  |
| 20% most deprived | 415 (24.4) | 367 (30.3) | 48 (9.8) |
| 20% to 40% | 405 (23.8) | 320 (26.4) | 85 (17.3) |
| 40% to 60% | 375 (22.0) | 261 (21.6) | 114 (23.3) |
| 60% to 80% | 293 (17.2) | 183 (15.1) | 110 (22.4) |
| 20% least deprived | 213 (12.5) | 80 (6.6) | 133 (27.1) |
| Not reported | 184 (9.8) | 156 (11.4) | 28 (5.4) |
| HBeAg status |  |  |  |
| Negative | 814 (66.3) | 541 (67.1) | 273 (64.7) |
| Positive | 414 (33.7) | 265 (32.9) | 149 (35.3) |
| Not available | 657 (34.9) | 561 (41.0) | 96 (18.5) |
| Anti-HBe status |  |  |  |
| Negative | 460 (35.5) | 305 (34.5) | 155 (37.5) |
| Positive | 836 (64.5) | 578 (65.5) | 258 (62.5) |
| Not available | 589 (31.2) | 484 (35.4) | 105 (20.3) |
| HBV VL, log10 IU/ml | 3.0 [1.5, 5.3] | 2.7 [1.5, 5.0] | 3.6 [1.9, 5.8] |
| HBV VL category |  |  |  |
| <20 IU/ml | 166 (8.8) | 120 (8.8) | 46 (8.9) |
| 20 - <2000 IU/ml | 842 (44.7) | 653 (47.8) | 189 (36.5) |
| 2000 - <20,000 IU/ml | 231 (12.3) | 155 (11.3) | 76 (14.7) |
| >=20,000 IU/ml | 646 (34.3) | 439 (32.1) | 207 (40.0) |
| ALT, IU/L | 35 [22, 58] | 34 [21, 57] | 37 [25, 60] |
| ALT, not available | 123 (6.5) | 87 (6.4) | 36 (6.9) |
| AST, IU/L | 33 [25, 48] | 32 [25, 46] | 35 [26, 52] |
| AST, not available | 848 (45.0) | 475 (34.7) | 373 (72.0) |
| Platelets, 10<sup>9</sup>/L | 203 [159, 246] | 201 [158, 246] | 208 [164, 246] |
| Platelets, not available | 247 (13.1) | 211 (15.4) | 36 (6.9) |
| Albumin, g/L | 40 [37, 44] | 41 [37, 44] | 40 [37, 44] |
| Albumin, not available | 236 (12.5) | 201 (14.7) | 35 (6.8) |
| ALP, IU/L | 78 [62, 101] | 76 [60, 96] | 85 [65, 142] |
| ALP, not available | 239 (12.7) | 204 (14.9) | 35 (6.8) |
| Bilirubin, µmol/L | 10 [7, 13] | 9 [7, 13] | 10 [7, 14] |
| Bilirubin, not available | 269 (14.3) | 92 (6.7) | 177 (34.2) |
| eGFR category |  |  |  |
| >=90 mL/min/1.73 m<sup>2</sup> | 679 (48.6) | 425 (44.0) | 254 (58.8) |
| >=60 & <=89 mL/min/1.73 m<sup>2</sup> | 569 (40.7) | 410 (42.5) | 159 (36.8) |
| <=59 mL/min/1.73 m<sup>2</sup> | 149 (10.7) | 130 (13.5) | 19 (4.4) |
| Not available | 488 (25.9) | 402 (29.4) | 86 (16.6) |
| Urea, mmol/L | 4.9 [4.0, 6.1] | 4.9 [3.9, 6.2] | 4.9 [4.1, 6.0] |
| Urea, not available | 183 (9.7) | 119 (8.7) | 64 (12.3) |
| Treatment regimens |  |  |  |
| TDF | 1048 (55.6) | 847 (62.0) | 201 (38.8) |
| ETV | 379 (20.1) | 231 (16.9) | 148 (28.6) |
| ETV+TDF | 150 (8.0) | 82 (6.0) | 68 (13.1) |
| LAM/ADE+TDF | 76 (4.0) | 45 (3.3) | 31 (6.0) |
| LAM/ADE+ETV | 51 (2.7) | 17 (1.2) | 34 (6.6) |
| Other regimens | 181 (9.6) | 145 (10.6) | 36 (6.9) |

*Data are the number (%) or median (IQR). HBV, Hepatitis B Virus; VL, viral load; NA, nucleos(tide) analogue; IMD, Index of Multiple Deprivation; ALT, alanine aminotransferase; TDF, tenofovir disoproxil fumarate; ETV, entecavir; LAM, lamivudine; ADE, adefovir.*

### Distinct classes of virologic trajectories in CHB patients identify heterogeneity in response to NA therapy

An overview of virologic trajectories in all individual patients on NA therapy is shown in **Figure S2**. With latent class mixed modelling in the derivation cohort, five mutually exclusive HBV DNA VL trajectories classes were identified (**Figure 1, Table S3**), as follows:

- Class 1 (N = 827, 60.5%) – ‘Long term suppression’;
- Class 2 (N = 254, 18.6%) – ‘Timely virological suppression’;
- Class 3 (N = 140, 10.2%) – ‘Persistent moderate viraemia’;
- Class 4 (N = 44, 3.2%) – ‘Persistent high-level viraemia’;
- Class 5 (N = 102, 7.5%) – ‘Slow virological suppression’.

**Figure 1.**
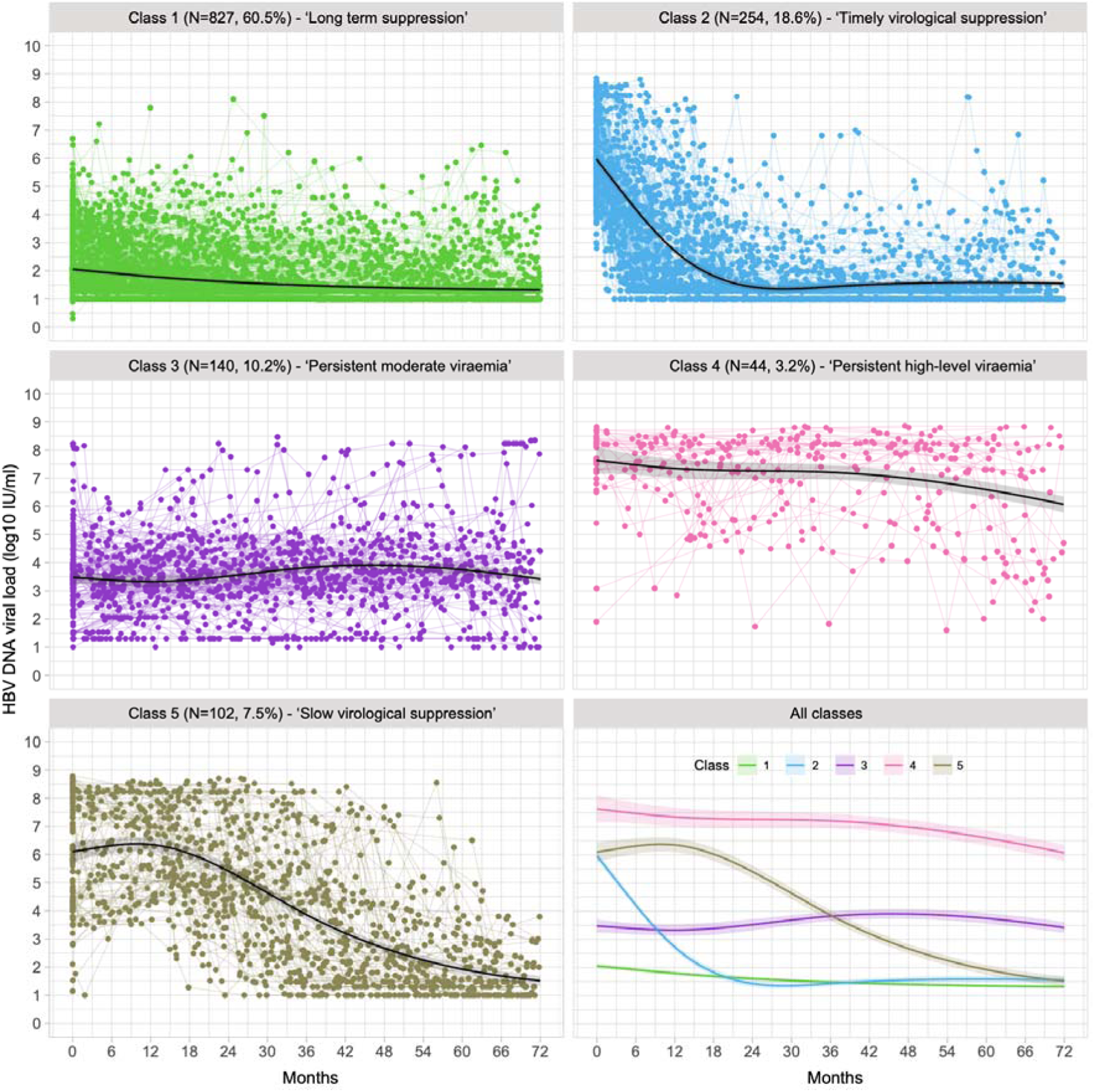
Individual trajectories of HBV VL for adults with chronic HBV infection on NA treatment divided into 5 classes using latent class mixed modelling. Data presented for derivation cohort (n=1367 individuals). Dots represent the raw values of VL linked by coloured lines for each individual, and solid black lines with shading area represent the predicted VL trajectory patterns with 95% confidence intervals. HBV, Hepatitis B Virus; VL, viral load; NA, nucleos/tide analogue.

The model showed a good discrimination of 0.93, with an entropy of 0.90, as well as high performance in classification with average posterior probability assignment (APPA) for each class of 0.9509, 0.8543, 0.9221, 0.9436, and 0.9330 (**Table S4**). In the majority of patients in class 2, VL suppressed to <50 IU/ml on treatment, which occurred at a median of 14 months (IQR: 9-23 months) while in class 5 this level of suppression was attained at a median of 51 months (IQR: 39-63 months). The trajectory seen in patients with long-term viral suppression (class 1) is mostly likely to represent later time points of follow up of the same group as those with suppressed VL from higher baseline (class 2), so these two groups largely reflect the same clinical and biological response, but with data captured at different stages of the treatment journey.

Using the latent class mixed model estimated from the derivation cohort, we then classified VL trajectories of patients in the external validation cohort into the five defined classes, which also showed good discrimination of 0.93 and entropy of 0.88 (**Table S4**). The observed VL trajectories of the validation cohort against the identified five classes is illustrated in **Figure S3.**

### Demographic and laboratory characteristics at baseline varied by VL class

Based on pooled data from derivation and validation cohorts, in 79.0% of longitudinal treatment episodes, there was an ‘**optimal**’ virological response (classes 1 and 2), while in 21% the virologic response was ‘**sub-optimal**’, i.e. incomplete, non-sustained, or delayed (classes 3, 4 and 5) (**Figure 2**).

**Figure 2.**
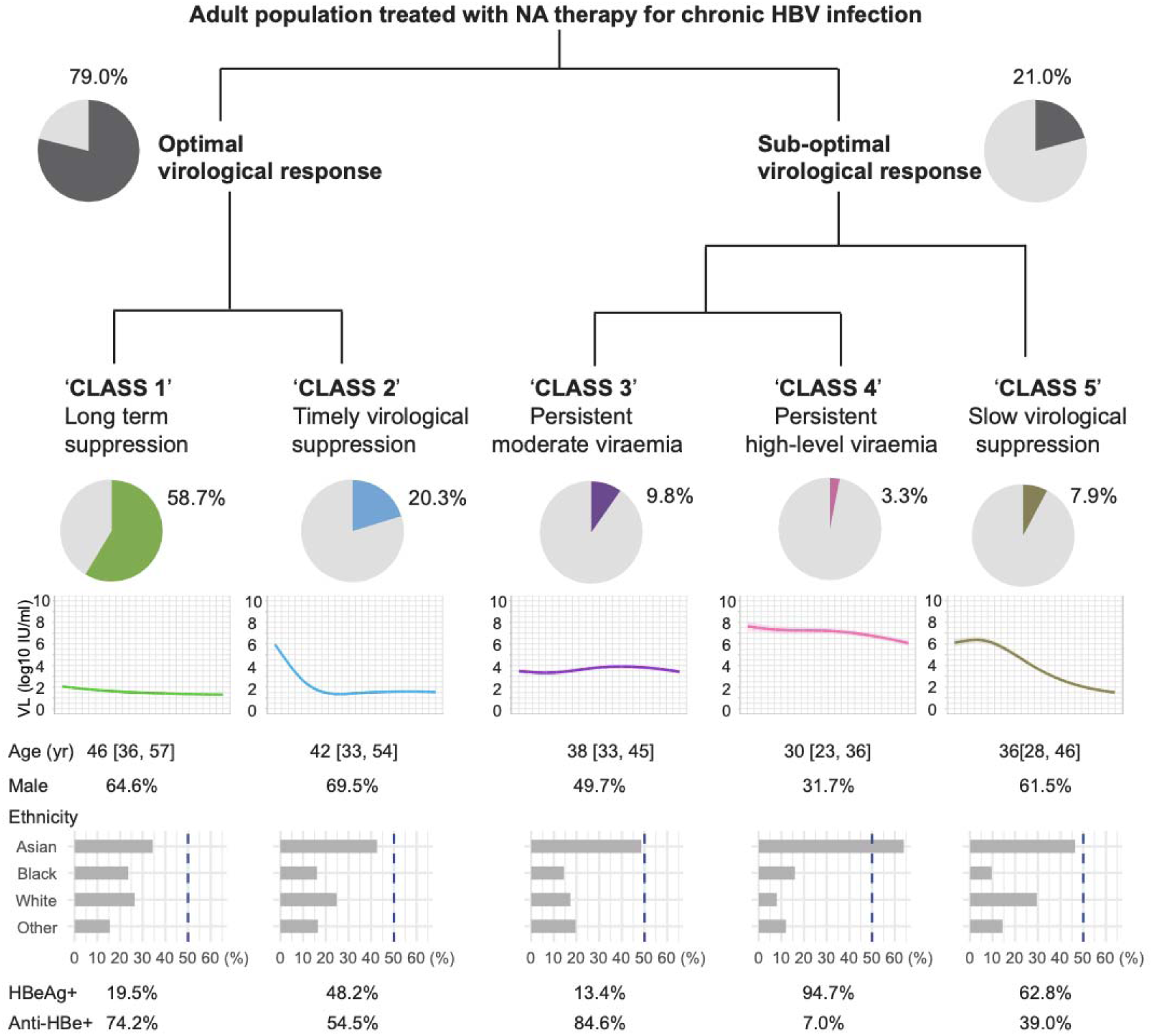
Schematic view of division of adult population with chronic HBV infection treated with NA therapy into five viral load classes. Data presented for 1885 individuals by pooling the derivation and validation cohorts. Proportion of the population in each class and characteristics of each class are shown. Age in years is given as median [interquartile range]. HBV, Hepatitis B Virus; NA, nucleos/tide analogue.

Baseline characteristics stratified by VL classes are presented in **Table 2** for the overall study cohort (and can be found in supplementary data for the derivation and validation cohorts separately, **Tables S5** and **Table S6** respectively), showing that individuals who were classified into different VL classes had significantly different characteristics at baseline. The majority of those in classes 1, 2 and 5 were male (64.6%, 69.5%, and 61.5% respectively), whereas class 3 had a balanced distribution of both sexes, and class 4 predominantly consisted of females (68.3%), p<0.001 (**Table 2**). Classes 3, 4 and 5 were younger than classes 1 and 2 (p<0.001), which may suggest the treatment duration was shorter for these patients.

**Table 2.** Characteristics of patients at presentation stratified by the virologic trajectory patterns identified by latent class mixed modelling in the derivation cohort (n=1885)

| Characteristics | Class 1<br>N =1106 (58.7%) | Class 2<br>N=383 (20.3%) | Class 3<br>N=185 (9.8%) | Class 4<br>N=63 (3.3%) | Class 5<br>N=148 (7.9%) | p value |
| --- | --- | --- | --- | --- | --- | --- |
|  | VL long term suppressed | VL suppressed timely | VL persistent with moderate levels | VL persistent with high levels | VL suppressed slowly |  |
| Sex, male | 715 (64.6) | 266 (69.5) | 92 (49.7) | 20 (31.7) | 91 (61.5) | <0.001 |
| Age, years | 46 [36, 57] | 42 [33, 54] | 38 [33, 45] | 30 [23, 36] | 36 [28, 46] | <0.001 |
| Age group, years |  |  |  |  |  | <0.001 |
| 18-24 | 19 (1.7) | 16 (4.2) | 5 (2.7) | 19 (30.2) | 18 (12.2) |  |
| 25-34 | 213 (19.3) | 99 (25.8) | 59 (31.9) | 25 (39.7) | 46 (31.1) |  |
| 35-44 | 274 (24.8) | 98 (25.6) | 71 (38.4) | 14 (22.2) | 41 (27.7) |  |
| 45-54 | 259 (23.4) | 84 (21.9) | 39 (21.1) | 3 (4.8) | 24 (16.2) |  |
| 55-64 | 196 (17.7) | 58 (15.1) | 10 (5.4) | 2 (3.2) | 18 (12.2) |  |
| 65-74 | 111 (10.0) | 18 (4.7) | 1 (0.5) | 0 (0.0) | 1 (0.7) |  |
| >=75 | 34 (3.1) | 10 (2.6) | 0 (0.0) | 0 (0.0) | 0 (0.0) |  |
| Ethnic group |  |  |  |  |  |  |
| Asian | 318 (34.4) | 132 (42.6) | 84 (48.6) | 32 (64.0) | 58 (46.4) | <0.001 |
| Black | 219 (23.7) | 50 (16.1) | 25 (14.5) | 8 (16.0) | 12 (9.6) |  |
| White | 245 (26.5) | 77 (24.8) | 30 (17.3) | 4 (8.0) | 37 (29.6) |  |
| Mixed/other ethnicity | 143 (15.5) | 51 (16.5) | 34 (19.7) | 6 (12.0) | 18 (14.4) |  |
| Not reported | 181 (16.4) | 73 (19.1) | 12 (6.5) | 13 (20.6) | 23 (15.5) |  |
| IMD (decile) | 5 [3, 7] | 5 [2, 7] | 5 [3, 7] | 4 [1, 7] | 5 [2, 7] | 0.425 |
| IMD category |  |  |  |  |  |  |
| 20% most deprived | 225 (22.0) | 96 (27.2) | 29 (22.1) | 20 (35.1) | 45 (33.1) |  |
| 20% to 40% | 277 (27.1) | 69 (19.5) | 28 (21.4) | 10 (17.5) | 21 (15.4) |  |
| 40% to 60% | 223 (21.8) | 76 (21.5) | 29 (22.1) | 12 (21.1) | 35 (25.7) |  |
| 60% to 80% | 177 (17.3) | 63 (17.8) | 27 (20.6) | 9 (15.8) | 17 (12.5) |  |
| 20% least deprived | 122 (11.9) | 49 (13.9) | 18 (13.7) | 6 (10.5) | 18 (13.2) |  |
| Not reported | 82 (7.4) | 30 (7.8) | 54 (29.2) | 6 (9.5) | 12 (8.1) |  |
| HBeAg status |  |  |  |  |  |  |
| Negative | 492 (80.5) | 158 (51.8) | 116 (86.6) | 3 (5.3) | 45 (37.2) | **<0.001** |
| Positive | 119 (19.5) | 147 (48.2) | 18 (13.4) | 54 (94.7) | 76 (62.8) |  |
| Not available | 495 (44.8) | 78 (20.4) | 51 (27.6) | 6 (9.5) | 27 (18.2) |  |
| Anti-HBe status |  |  |  |  |  |  |
| Negative | 176 (25.8) | 138 (45.5) | 21 (15.4) | 53 (93.0) | 72 (61.0) | **<0.001** |
| Positive | 506 (74.2) | 165 (54.5) | 115 (84.6) | 4 (7.0) | 46 (39.0) |  |
| Not available | 424 (38.3) | 80 (20.9) | 49 (26.5) | 6 (9.5) | 30 (20.3) |  |
| HBV VL, log10 IU/ml | 1.5 [1.3, 2.6] | 6.3 [5.1, 7.7] | 3.7 [2.8, 4.5] | 8.1 [7.5, 8.4] | 6.3 [4.6, 8.2] | **<0.001** |
| HBV VL categories, IU/ml |  |  |  |  |  | **<0.001** |
| <20 | 164 (14.8) | 0 (0.0) | 1 (0.5) | 0 (0.0) | 1 (0.7) |  |
| 20 - <2000 | 763 (69.0) | 4 (1.0) | 65 (35.1) | 2 (3.2) | 8 (5.4) |  |
| 2000 - <20,000 | 121 (10.9) | 29 (7.6) | 68 (36.8) | 0 (0.0) | 13 (8.8) |  |
| >=20,000 | 58 (5.2) | 350 (91.4) | 51 (27.6) | 61 (96.8) | 126 (85.1) |  |
| ALT, IU/L | 29 [20, 47] | 64 [37, 125] | 32 [21, 41] | 35 [22, 50] | 41 [32, 69] | **<0.001** |
| ALT, not available | 41 (3.7) | 24 (6.3) | 21 (11.4) | 12 (19.0) | 25 (16.9) |  |
| AST, IU/L | 31 [24, 41] | 45 [32, 74] | 28 [23, 33] | 27 [23, 57] | 33 [27, 60] | **<0.001** |
| AST, not available | 467 (42.2) | 147 (38.4) | 106 (57.3) | 42 (66.7) | 86 (58.1) |  |
| Platelets, 10<sup>9</sup>/L | 199 [156, 248] | 201 [158, 238] | 220 [170, 256] | 206 [179, 244] | 213 [179, 262] | **0.001** |
| Platelets, not available | 83 (7.5) | 54 (14.1) | 40 (21.6) | 25 (39.7) | 45 (30.4) |  |
| Albumin, g/L | 40 [36, 43] | 40 [37, 43] | 43 [40, 46] | 41 [37, 45] | 41 [38, 44] | **<0.001** |
| Albumin, not available | 81 (7.3) | 53 (13.8) | 35 (18.9) | 25 (39.7) | 42 (28.4) |  |
| ALP, IU/L | 79 [62, 101] | 82 [63, 108] | 69 [55, 96] | 74 [61, 101] | 75 [61, 102] | **0.005** |
| ALP, not available | 81 (7.3) | 53 (13.8) | 35 (18.9) | 25 (39.7) | 45 (30.4) |  |
| Bilirubin, µmol/L | 10 [7, 13] | 10 [7, 14] | 9 [6, 12] | 8 [6, 11] | 10 [7, 13] | **0.001** |
| Bilirubin, not available | 109 (9.9) | 60 (15.7) | 38 (20.5) | 20 (31.7) | 42 (28.4) |  |
| eGFR categories,<br>mL/min/1.73 m<sup>2</sup> |  |  |  |  |  | **<0.001** |
| >=90 | 397 (45.6) | 147 (51.6) | 66 (51.2) | 17 (77.3) | 52 (57.1) |  |
| >=60 & <=89 | 350 (40.2) | 125 (43.9) | 54 (41.9) | 5 (22.7) | 35 (38.5) |  |
| <=59 | 123 (14.1) | 13 (4.6) | 9 (7.0) | 0 (0.0) | 4 (4.4) |  |
| Not available | 236 (21.3) | 98 (25.6) | 56 (30.3) | 41 (65.1) | 57 (38.5) |  |
| Urea, mmol/L | 5.0 [4.0, 6.4] | 4.8 [4.0, 5.8] | 4.6 [3.9, 5.8] | 4.0 [3.3, 4.6] | 5.0 [4.1, 5.9] | **<0.001** |
| Urea, not available | 61 (5.5) | 29 (7.6) | 38 (20.5) | 19 (30.1) | 36 (24.3) |  |
| Treatment regimens |  |  |  |  |  | **<0.001** |
| TDF | 562 (50.8) | 253 (66.1) | 115 (62.2) | 40 (63.5) | 78 (52.7) |  |
| ETV | 234 (21.2) | 68 (17.8) | 40 (21.6) | 8 (12.7) | 29 (19.6) |  |
| ETV+TDF | 64 (5.8) | 38 (9.9) | 17 (9.2) | 9 (14.3) | 22 (14.9) |  |
| LAM/ADE+TDF | 58 (5.2) | 8 (2.1) | 3 (1.6) | 1 (1.6) | 6 (4.1) |  |
| LAM/ADE+ETV | 47 (4.2) | 1 (0.3) | 2 (1.1) | 0 (0.0) | 1 (0.7) |  |
| Other regimens | 141 (12.7) | 15 (3.9) | 8 (4.3) | 5 (7.9) | 12 (8.1) |  |

Data are the number (%) or median [IQR]. IMD, Index of Multiple Deprivation; VL, viral load; ALT, alanine aminotransferase; TDF, tenofovir disoproxil fumarate; ETV, entecavir; LAM, lamivudine; ADE, adefovir. Comparison was conducted across non-missing categories for a categorical variable.

Class 4 contained a higher proportion of individuals of Asian ancestry (64.0%) compared to other groups (34.4% to 48.6%, p<0.001). Most patients in class 3 (VL persistent at moderate levels) were anti-HBe positive (84.6%), while 94.7% in class 4 were HBeAg positive at baseline, which may account for differences in VL levels. Patients in class 2 had a significantly higher median ALT level (median 64, [IQR 37, 125] IU/L) at baseline, followed by class 5 (median 41, [32, 69] IU/L), compared to other classes (29 [20, 47] IU/L for class 1, 32 [21, 41] IU/L for class 3, 35 [22, 50] IU/L for class 4), p<0.001.

### ALT decreased over time in classes with suppressed VL

We assessed longitudinal trends of ALT by linear mixed effects models (considering repeated measurements, and adjusted for age, sex, and ethnicity). ALT decreased at differing rates over time in the VL suppressed groups (classes 1, 2, and 5; all p <0.001), in line with declining VL trajectories of these classes, whilst ALT levels of patients in classes 3 and 4 (with persistent viraemia) did not significantly decrease over time or worsened (**Figure S4**, **Table 3**). In class 1, ALT levels continuously decreased but only marginally so (coefficient: -0.0026, 95% CI: -0.0034 to -0.0018), as most of the patients already had normalised ALT at baseline compared to other classes (**Figure S5**).

**Table 3.** Longitudinal trend of ALT over time for distinct virologic trajectory patterns for patients with chronic HBV infection on NA therapy in the overall study cohort.

| | Intercept (95% CI),<br>log IU/L | Coefficient $\beta$ (95% CI) |
| --- | --- | --- |
| Class 1 (VL long term suppressed) | 3.95 (3.85, 4.05) *** | -0.0026 (-0.0034, -0.0018) *** |
| Class 2 (VL suppressed timely) | 4.39 (4.29, 4.50) *** | -0.0108 (-0.0122, -0.0094) *** |
| Class 3 (VL persistent with moderate levels) | 4.01 (3.88, 4.13) *** | -0.0007 (-0.0026, 0.0012) |
| Class 4 (VL persistent with high levels) | 4.13 (3.96, 4.30) *** | -0.0006 (-0.0037, 0.0026) |
| Class 5 (VL suppressed slowly) | 4.48 (4.35, 4.60) *** | -0.0085 (-0.0106, -0.0065) *** |

*Derivation and validation cohorts were combined according to the VL trajectories. 1883 patients (out of 1885, 99.89%) had longitudinal measurements of ALT for modelling. The longitudinal trends of ALT over time were assessed by linear mixed effects models with random intercepts and slopes. All linear mixed effects models were adjusted for demographics (including age, sex, and ethnicity). \*\*\* indicates p value <0.001. HBV, Hepatitis B Virus; VL, viral load; NA, nucleos(tide) analogue; ALT, alanine aminotransferase.*

We compared changes in ALT in class 5 compared to class 2, because they had similar baseline ALT and VL levels. ALT levels in class 5 decreased more slowly compared to class 2 (Coefficient: -0.0108, 95% CI: -0.0122 to -0.0094 for class 2 vs -0.0085, 95% CI: -0.0106 to -0.0065 for class 5, **Table 3**, **Figure S4**). Varying proportions of patients had elevated ALT over time among different classes (**Figure S5-S6**). Class 1 maintained similar ALT levels over time, class 2 experienced timely improvements of ALT levels, and class 5 showed a slower improvement, in keeping with VL suppression trajectories.

### Risk of liver fibrosis/cirrhosis varied by VL class

For disease progression analysis, we analysed 1412 individuals for whom data were available on fibrosis/cirrhosis status, combining derivation and validation cohorts. Overall, those with liver fibrosis or cirrhosis compared to those without were more likely to be male (71.4% vs 58.4%) and older (46 years vs 40 years) (p<0.001). Individuals in class 5 (who suppressed VL slowly) were more likely to have liver fibrosis/cirrhosis compared to other classes (p<0.001) (**Table 4**).

**Table 4.**
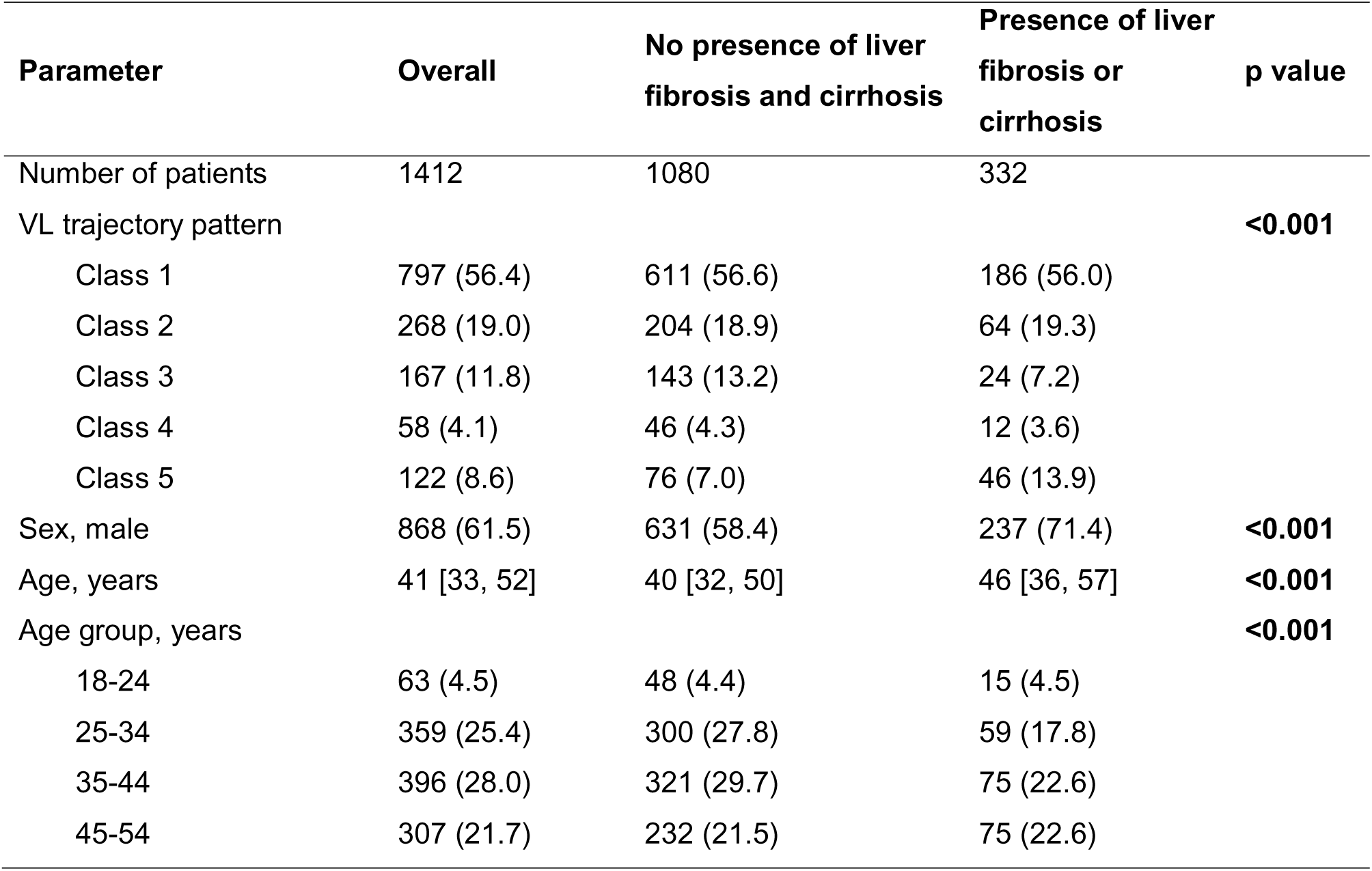
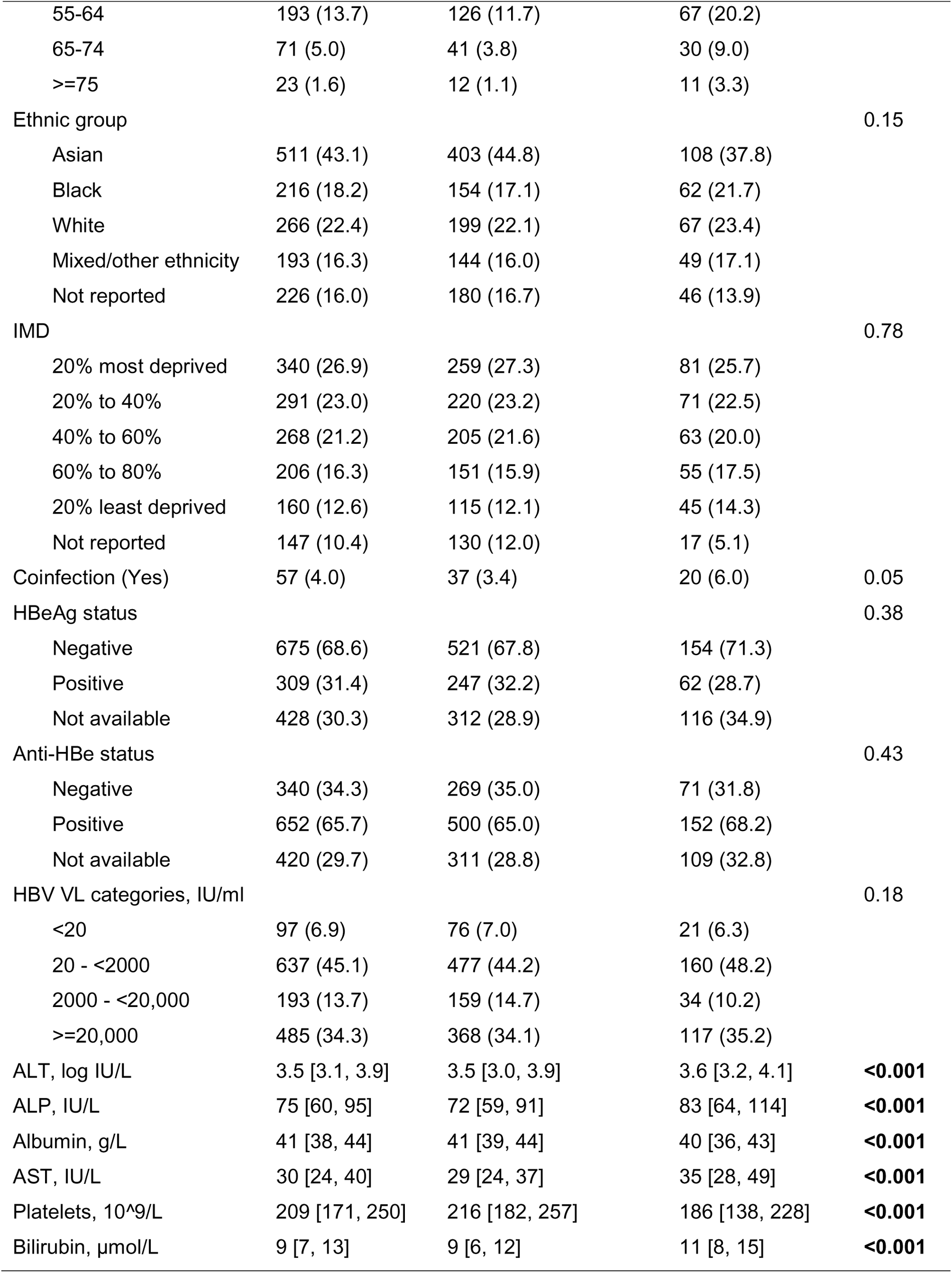
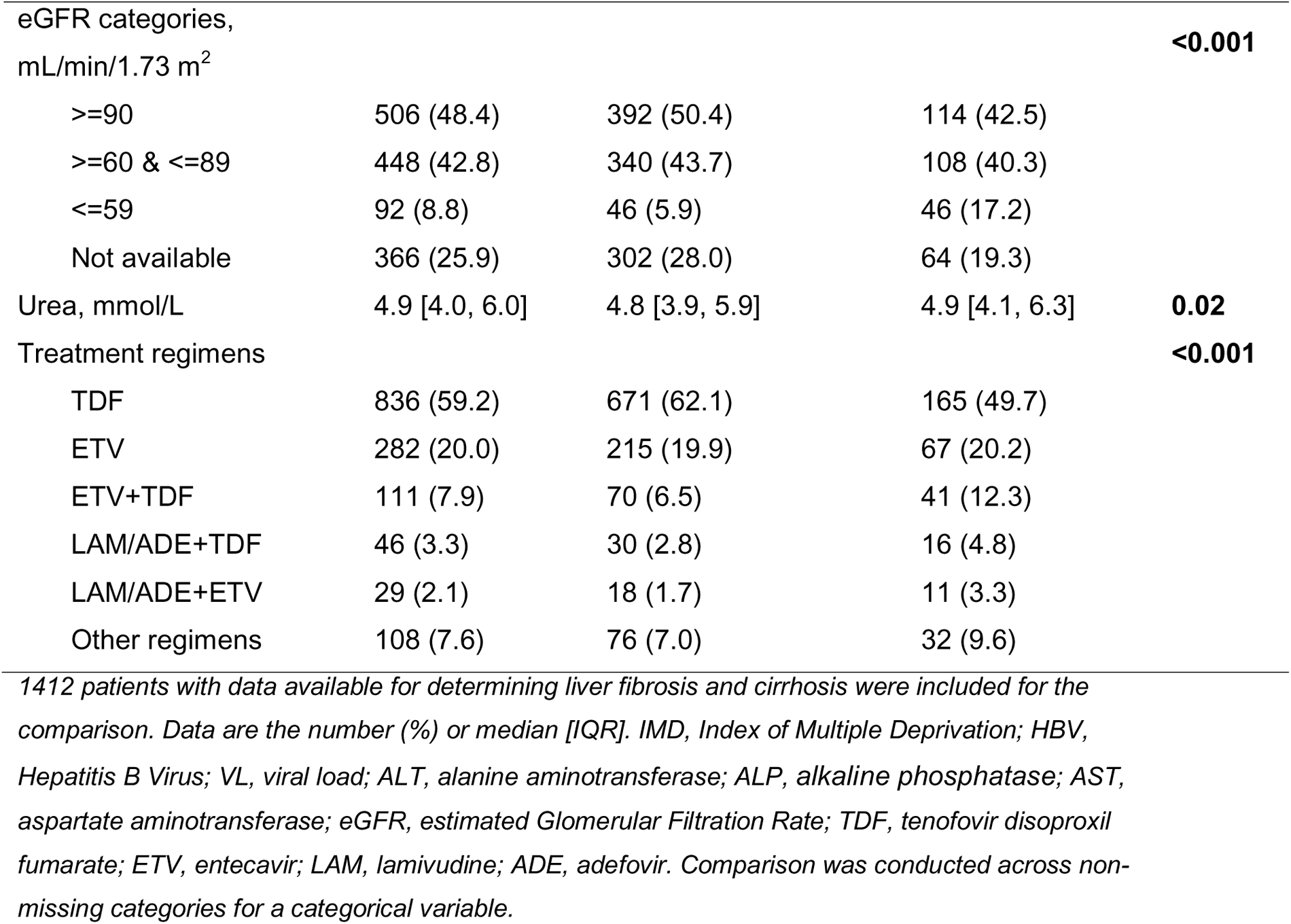
Baseline characteristics of patients with vs without the presence of liver fibrosis and/or cirrhosis in the overall cohorts, combining derivation and validation cohorts according to the VL trajectory classes.

Kaplan-Meier (K-M) analysis demonstrated that individuals in class 5 (slow VL suppression) were more likely to have liver fibrosis/cirrhosis over follow-up to 108 months, compared to class 1 (p=0.003, **Figure 3**). The cumulative probability of being free of fibrosis/cirrhosis was similar between class 1 and class 2 (p=0.73), as well as between classes 3 and class 4 (p=0.47) (**Figure 3**). Individuals in class 3 were less likely to have liver fibrosis and cirrhosis at baseline compared to class 1 but the cumulative probability of being free of fibrosis/cirrhosis for class 3 decreased rapidly over time, which may reflect increasing age (**Figure 3**). A similar trend was observed for class 4 (**Figure 3**, **Table S7**). K-M curves were constructed without adjustment for confounders, so other attributes of the class (age, sex, HBeAg status) may be driving the observed differences.

**Figure 3.**
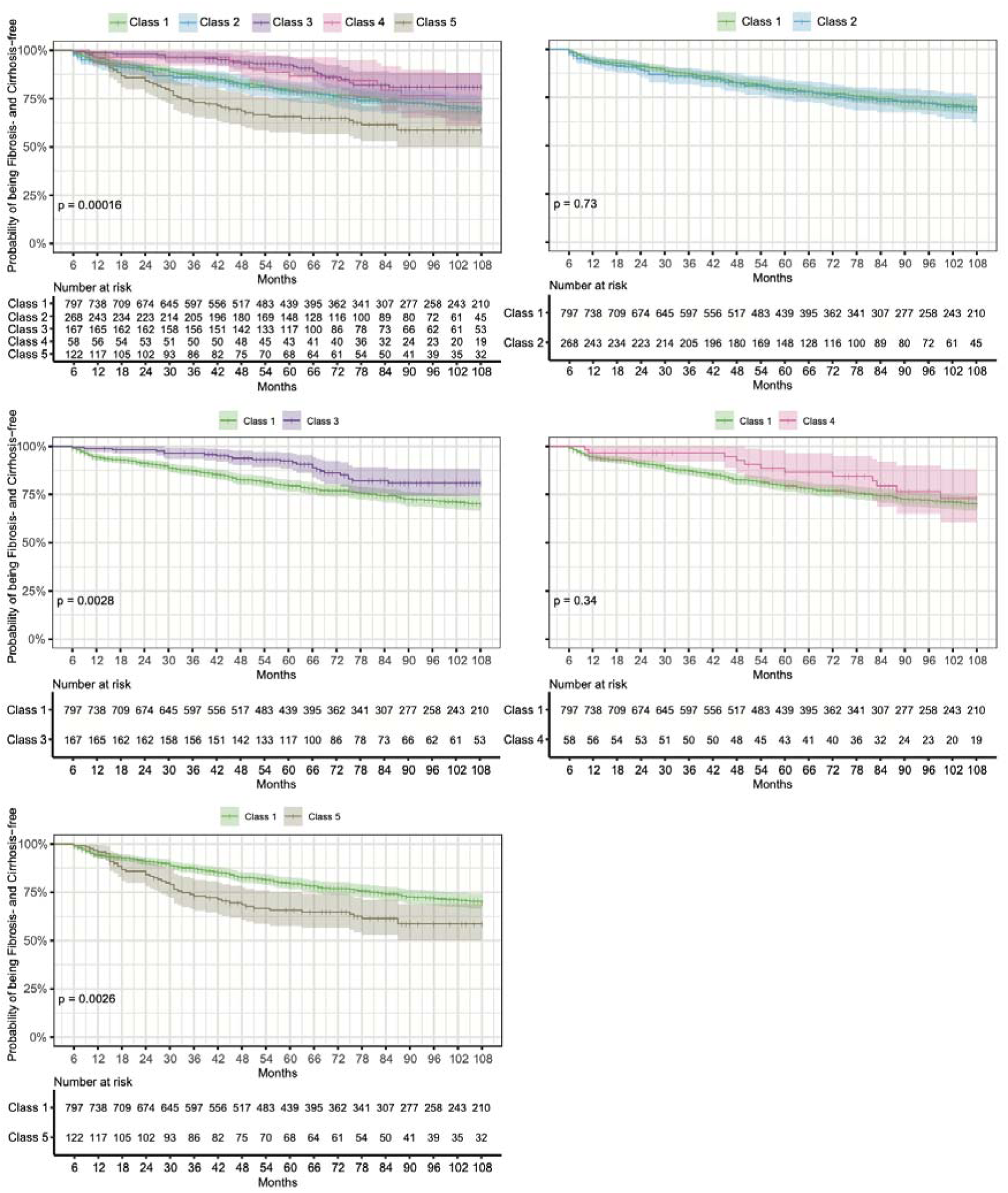
Progression to liver fibrosis and/or cirrhosis and VL trajectories of adult population with chronic HBV infection treated with NA therapy plotted as Kaplan-Meier (K-M) curves. K-M curves were truncated at 75% quantiles of follow up duration (months=108) of liver fibrosis/cirrhosis status. Shading areas represent 95% confidence intervals. The shape “|” indicates censoring. Note that classes 3 and 4 are characterised by being more female and younger (see Figure 2), which may be associated with slower rate of progression. HBV, Hepatitis B Virus; VL, viral load; NA, nucleos/tide analogue.

Multivariate analysis (fully adjusted for demographics, laboratory parameters, and treatment regimens) showed that patients in class 5 had ∼2-fold increased hazards of progression to fibrosis/cirrhosis compared to those patients with VL long-term suppressed (class 1) (adjusted HR, 2.24; 95% CI, 1.55-3.24) (**Figure 4, Table S8**). Compared to those patients aged 25-34 years, those aged 55-64 years, 65-74 years, and ≥75 years had ∼2-fold (adjusted HR 1.95; 95% CI, 1.33-2.85), ∼3-fold (adjusted HR, 3.13; 95% CI, 1.93-5.07), and ∼6-fold (adjusted HR, 5.70; 95% CI, 2.84-11.44) increased hazards of progression to liver fibrosis/cirrhosis, respectively, while males had 1.40-fold increased hazards of progression (adjusted HR, 1.40; 95% CI, 1.06-1.84) compared to females. As expected, lower albumin and platelets, and higher AST and ALP, were significantly associated with fibrosis/cirrhosis (**Figure 4, Table S8**). Sensitivity analysis only adjusting for age and sex, shows similar hazards ratio for each VL trajectory class (**Table S9**).

**Figure 4.**
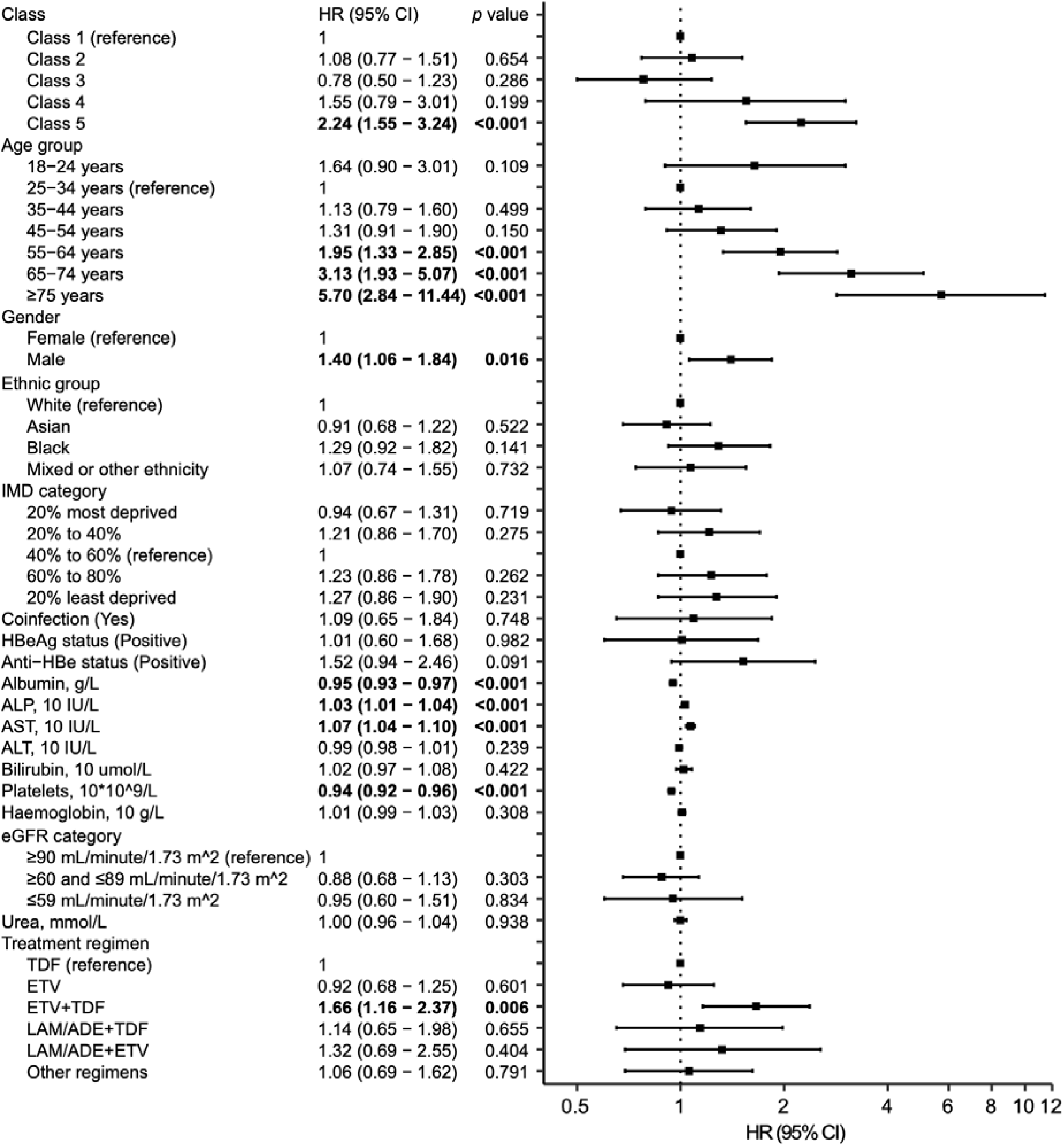
Multivariate Cox proportional-hazards model investigating associations of different VL trajectory patterns with liver disease progression to fibrosis and/or cirrhosis among adults with chronic HBV infection on NA therapy. Forest plot showing the hazard ratios and 95% confidence intervals (CIs) for the development of liver fibrosis and cirrhosis. HBV, Hepatitis B Virus; VL, viral load; NA, nucleos/tide analogue.

ROC analyses demonstrated that the predictive ability of VL trajectory for liver fibrosis and cirrhosis (AUC=0.66) was comparable to that of other well-known predictors in previous studies, such as age (AUC=0.65), sex (AUC=0.63), and platelets (0.67) (**Figure 5A**; all pairwise p values non-significant, **Table S10**). Furthermore, we demonstrated the addition of VL trajectory into the combinations of other predictors for liver fibrosis and cirrhosis significantly improved the prediction performance (**Figure 5B**, **Table S11,** all p values <0.01), which can further improve early identification of patients at risk of liver complications.

**Figure 5.**
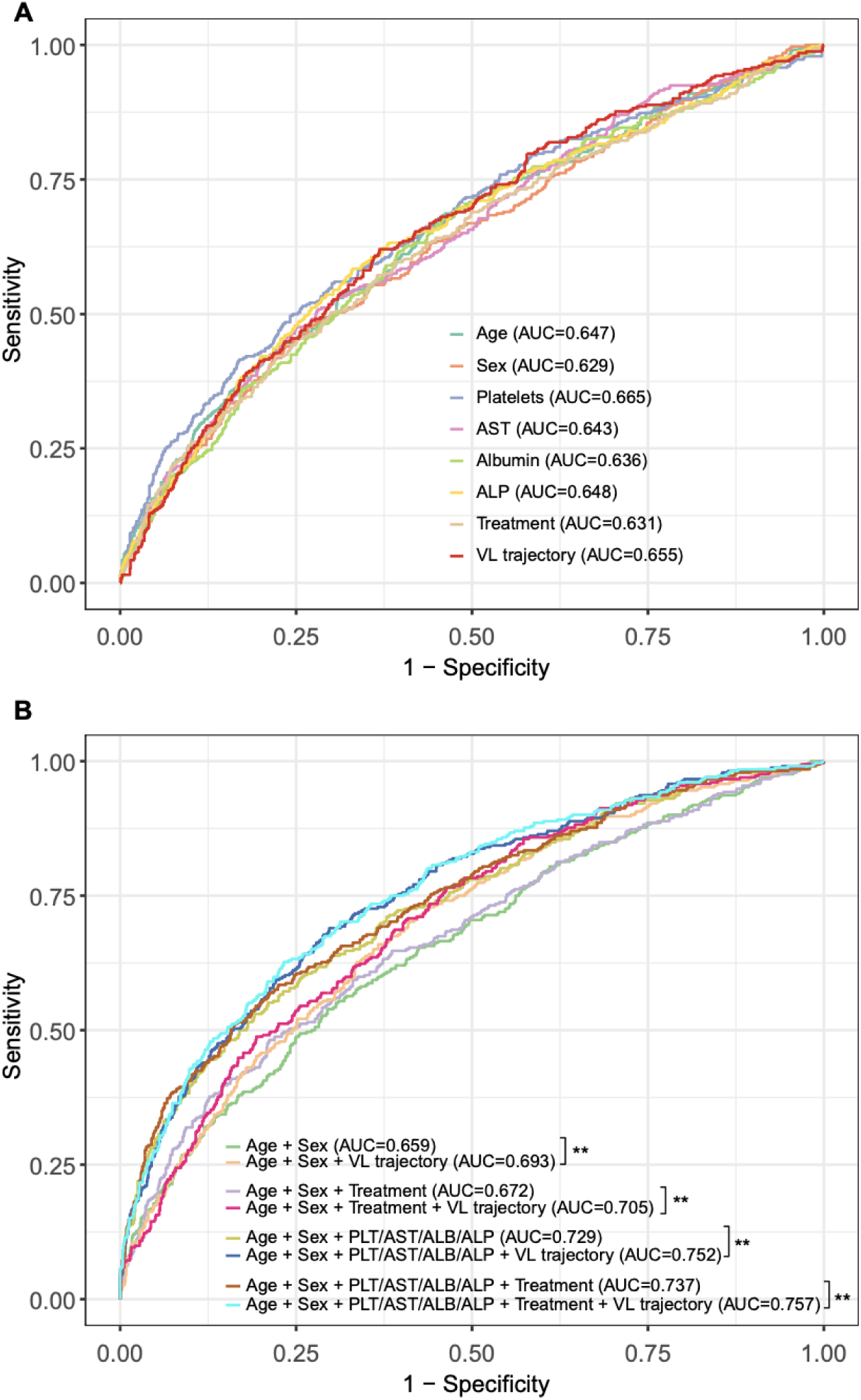
ROC curves of HBV VL trajectory and other identified predictors for liver fibrosis and cirrhosis: (A) single identified predictor (B) combination of different identified predictors. All models were adjusted for other parameters that were included in multivariate analysis. ** in panel B indicates p value <0.01. ROC, receiver operating characteristic curve; VL, viral load. HBV, Hepatitis B Virus; VL, viral load; NA, nucleos/tide analogue.

### Minimum number of repeated measurements of VL required to assign class

We conducted sensitivity analysis on the validation cohort, in order to ascertain how robustly individuals can be classified into one of our five classes with a limited number of repeated VL measurements. Using only the first two VL measurements per individual (but retaining the requirement for ≥6 month follow-up measurements of HBV DNA VL in line with our original eligibility criteria), the classification still achieved a good discrimination (0.83) and entropy (0.73). These data spanned a median follow up duration of 12 months (IQR: 7-19) (**Table S12**).

We repeated this analysis, but this time removing restrictions on the minimum follow up duration for each patient, which demonstrated that ≥3 VL measurements are required for assigning an individual into one of our five classes with good discrimination (0.82) and entropy (0.72) (**Table S13**). These three measurements spanned a median of 12 months follow up (IQR: 6-21), similar to that in the above scenario. Therefore, individuals with 2 or 3 VL repeated measurements spanning a one-year period can be reliably assigned into a class based on this model.

### Relationship between VL classes and EASL definition of virological breakthrough

Based on definitions in EASL guidelines (6), we determined the proportion of individuals in our overall cohort, and in each of the five classes, who experienced virological breakthrough on NA treatment. We found breakthrough in 6.4% (120/1885), and the proportion in classes 1-5 was 4.0%, 2.9%, 17.3%, 30.2%, and 9.5%, respectively (**Supplementary Table S14**).

## Discussion

### Summary of novelty and key findings

We applied an integrated data pipeline to capture large scale, routinely collected, clinical and laboratory data to assess the longitudinal VL trajectory alongside clinical parameters in CHB patients from multiple secondary care centres across England. We identified five novel and distinct VL trajectory classes using an unbiased discovery method, and confirmed these using a validation set. Individuals assigned to these classes had distinct clinical, laboratory and demographic characteristics.

These real-world data are an important addition to our understanding of treatment responses, in contrast to much of the existing HBV treatment data which are derived from closely monitored and highly selected cohorts taking part in clinical trials. We are thus uniquely positioned to determine the outcomes of treatment across wide populations where diverse and complex factors operate to influence outcomes, including for example co-morbidity, poly-pharmacy, alcohol use, socioeconomic factors, and adherence to therapy. To our knowledge, this is the first study to comprehensively document and classify long-term virologic trajectories in a large and unselected population on HBV treatment, and to explore VL trajectory sub-cohorts/classes where virological response to treatment may be sub-optimal. Understanding VL suppression has implications for both individual disease progression and also HBV outcomes and transmission at a population level. As guidelines for the management of HBV infection are reviewed and may be relaxed over time, higher numbers of people will become eligible for NA treatment and there is a heightened need to understand responses and outcomes of therapy, and to provide resources and an evidence base to intervene when an adequate virologic response is not achieved.

In the majority of individuals (∼80%) HBV VL suppressed on treatment, assigned as classes 1 and 2. However, importantly, ∼20% of individuals treated with NA agents for CHB experience suboptimal virological responses, with persistent moderate or high VL (classes 3 and 4), or slow suppression of VL (class 5), which may have significant implications for clinical outcomes. Documenting and classifying these distinct on-treatment phenotypes can allow discrimination between satisfactory virologic treatment responses, versus situations in which a suboptimal response might expose an individual to excess clinical risks. In the latter situation, patients may benefit from enhanced monitoring and/or alterations to therapy. At a public health level, quantifying the risks of non-suppression of viraemia on NA treatment is important to allow modelling of population outcomes as increased numbers of people will be diagnosed and treatment eligible over the years ahead. For example, many new HBV diagnoses are being made as a result of the NHS England programme to implement routine ‘opt-out’ testing in Emergency Department attenders (33).

### Relationship between VL class and clinical outcome

Individuals in class 5 (slow suppression of VL) had an increased risk of liver fibrosis/cirrhosis within the observation period, compared to class 1. However, we did not find a significant association between non-suppressing groups (classes 3 and 4) and the risk of liver fibrosis/cirrhosis, potentially reflecting the younger age of these groups and enrichment for female sex which is known to be protective (34), compared to class 1. In keeping with this, the lower hazards of liver fibrosis/cirrhosis in class 3 were not significant and class 4 (persistent high viraemia) is close to significance after adjustment for the relevant confounders. In addition, the K-M analysis shows that, although patients in classes 3 and 4 at the early time points of observation were less likely to develop liver fibrosis/cirrhosis compared to class 1, they progressed to liver fibrosis/cirrhosis rapidly over time with increasing age. This highlights the need for longer follow up to determine associations with disease progression, which may only become apparent in older adults. Furthermore, increased sample size for classes 3 and 4 is needed to support further investigation into the association with liver disease progression.

After correction for confounders, only class 5 retained a significant relationship with the development of fibrosis/cirrhosis. This observation is not simply related to VL at baseline, as classes 2, 4 and 5 all started their trajectory with high VL, and univariate analysis showed that baseline VL was not significantly associated with disease progression (Table S8). We also conducted sensitivity analysis with a simpler model only adjusting for age and sex, showing similar results (class 5 associated with liver fibrosis/cirrhosis).

To determine the applicability of this approach to other real-world data, we evaluated the performance of the model using smaller number of VL measurements per patient, which demonstrated three VL measurements are required without restriction on minimum follow up duration, or two VL measurements at least six months apart, to discriminate between classes. Our model can thus be considered for predicting outcomes in different settings:

i. To identify individuals who are at high risk of non-suppression on NA treatment, and thus may warrant closer scrutiny to avoid complications;
ii. To improve the early prediction of liver fibrosis and cirrhosis, by incorporating VL trajectory classifications into existing scoring tools;
iii. To inform preventive interventions, exemplified by antiviral prophylaxis for the prevention of mother to child transmission (PMTCT), when a consistent and timely reduction in viraemia is required;
iv. To inform public health and programmatic interventions which seek to deliver on population elimination targets.

### Caveats and limitations

Our analysis was limited by the missingness of certain data. For example, we were unable to adjust for metabolic laboratory parameters such as total cholesterol, triglycerides and glucose, although emerging evidence suggests the combination of metabolic disease and CHB can accelerate the progression of liver pathology (35,36). As body mass index and alcohol were missing for majority of the study subjects in this study, we were unable to consider these risk factors in the analysis for the development of liver fibrosis and cirrhosis. Quantitative HBsAg (qHBsAg) levels are important for predicting immunological control and functional cure, however, qHBsAg was not routinely tested by all our sites, so we did not have sufficient data to analyse this variable.

The dataset does not contain details of the potential underlying causes for non-suppression of viraemia, which include treatment interruptions or incomplete adherence to therapy, inadequate drug levels, and genotypic viral resistance; further work is needed to dissect these causes and to understand their relative contributions. Our groups partly overlap with classification presented in EASL guidelines as we were able to identify individuals with virological breakthrough based on existing definitions. However, lack of robust treatment histories that include a documented treatment start date meant we could not reliably assign individuals to other EASL classes. Longer periods of follow up in large datasets are required to develop a better understanding of the association between non-suppressed VL patterns and outcomes, particularly risks of cirrhosis, HCC and transmission events. Prospectively, data collection through NIHR HIC network will continue, providing us with more power to identify significant long-term outcomes.

### Conclusions

There is significant heterogeneity in virologic response to HBV NA treatment. Complete virologic suppression on NA treatment can be slow or incomplete, correlating in some cases with enhanced clinical risk. Enhanced understanding of treatment response can be used to inform better risk-stratification, improved patient-centric clinical care, models of treatment response at a population level, and as a foundation to understand the additional impact of novel therapies as these become available. Additional scrutiny is needed to understand the determinants of non-suppression so that risk factors can be tackled. Optimising use of NA therapy is fundamental to reducing morbidity and mortality, and decreasing transmission, which are crucial attributes of international targets to eliminate HBV infection as a public health threat.

## Data Availability

All data produced in the present work are contained in the manuscript.

## Ethics approval

The research database for the NIHR HIC Viral Hepatitis and Liver Disease theme was approved by South Central—Oxford C Research Ethics Committee (REF Number: 21/SC/0060). All methods for data collection, transmission, and management for this study were carried out in accordance with relevant guidelines and regulations. The requirement for written informed consent was waived by South Central—Oxford C Research Ethics Committee, because data have been in effect anonymised before transfer to the research database.

## Author contributions

EB, PCM, and TW contributed to the conception and design of the work. EB, PCM, GSC, WG, SIK, EN, KW, and JD directed the data collation and implementation of the study. TW, CC, AJS, ST, KM, AF, JJ, BG, DP, LM, CRJ, HS, GR, SL, TN, KAV, CD, AIH, MG, FB, LE, LR, and DR contributed to the methodology development and/or the data acquisition, processing, interpretation, and management of study data, with the support from the NIHR HIC Viral Hepatitis and Liver Disease Consortium. TW and PCM designed the analytical strategy, and TW conducted the analysis, supervised by EB and PCM. PCM, EB, AJS, WG, SIK, GSC, and EN helped interpret the findings. TW and PCM wrote the original draft. All authors reviewed and revised the manuscript critically and approved the final version for publication.

## Acknowledgements

The authors thank all the research nurses and research administrative staff at the contributing sites including Cambridge University Hospitals NHS Foundation Trust, Imperial College Healthcare NHS Trust, Liverpool University Hospitals NHS Foundation Trust, Oxford University Hospitals NHS Foundation Trust, Southampton NHS Foundation Trust and University College London Hospitals NHS Foundation Trust for their help in data collection and submission, as well as Dr. Deepak Suri and Dr. Stuart Flanagan from the Department of Hepatology, UCLH for their advice and support of the data collection. The authors thank all Scientific Steering Committee members of the NIHR HIC Viral Hepatitis and Liver Disease theme for their support and approval of this study. We also thank the Oxford University Hospitals NHS Foundation Trust Clinical Data Warehouse team for providing the computing environment for this study and Charles Crichton and Jaimie Withers for ongoing management and support of the environment.

