## Supplemental figures and tables for "Classification of virologic trajectories during nucleos/tide analogue treatment of hepatitis B virus (HBV) infection"

**during nucleos/tide analogue treatment**

**of hepatitis B virus (HBV) infection**

Eleanor Barnes, Philippa C Matthews, William Gelson, Graham S Cooke, Salim I Khakoo, Eleni Nastouli, Jim Davies, Kerrie Woods, Alexander J Stockdale, Stephen Ryder, Ahmed Elsharkawy, Nicholas Easom, William Bernal, Shazaad Ahmad, Douglas Macdonald, Tingyan Wang, Cori Campbell, Cedric Tan, Simon Stanworth, Suzanne Maynard, Gail Roadknight, Stephanie Little, Kinga A Várnai, Ben Glampson, Dimitri Papadimitriou, Luca Mercuri, Christopher R Jones, Jakub Jaworski, Cai Davis, Florina Borca, Ashley Heinson, Michael George, Heidi MacNaughton, Yun Kim, Josune Olza Meneses, Louise English, Timothy Roberts, Luis Romão, David Ramlakhan, Stacy Todd, Heather Rogers, Karl McIntyre, Andrew Frankland, Hizni Salih, Theresa Noble, Lara Roberts, Finola Higgins, Javier Vilar, Ruth Norris, George Tilston, Ilina Serafimova, Sarah Montague, Juliette Verheyden, Irene Juurlink, Kathryn Jack, Alex Waldren-Glenn, Lizzie Poole, Victoria Day, Berit Reglar.

^†^ Joint last authors, represents equal contribution.

**Corresponding authors**: Professor Eleanor Barnes, The Peter Medawar Building for Pathogen Research, South Parks Road, Oxford, OX1 3SY, UK. Telephone: 01865 281547.

Professor Philippa C Matthews, The Francis Crick Institute, 1 Midland Road, London, NW1 1AT, UK.

**Key words**: HBV, viral load, longitudinal virological trajectories, liver fibrosis, cirrhosis, antiviral treatment, latent class mixed models, National Institute for Health and Care Research Health Informatics Collaborative (NIHR HIC)

**Word count**: 5253 words

**Number of figures and tables**: 5 figures, 4 tables

**ORCIDs:**

T.W – 0000-0002-8351-9494

C.C – 0000-0001-5890-7105

H.S – 0000-0002-5283-7799

K.A.V – 0000-0001-5695-9497

K.W – 0000-0001-8106-8290

J.D – 0000-0003-4664-6862

P.C.M – 0000-0002-4036-4269

E.B – 0000-0002-0860-0831

A.S – 0000-0002-5828-3328

C.D - 0000-0003-3710-1879

A.I.H - 0000-0001-8695-6203

### Supplementary Materials

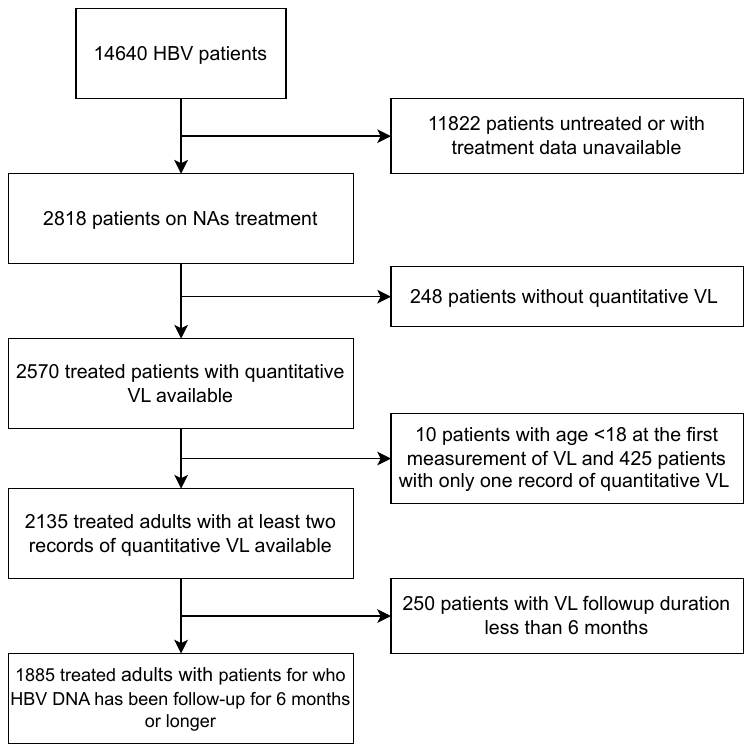

**Supplementary Figure S1.** Flowchart of patient selection for a study of viral load trajectories in adults receiving NA therapy for chronic HBV infection in the UK.

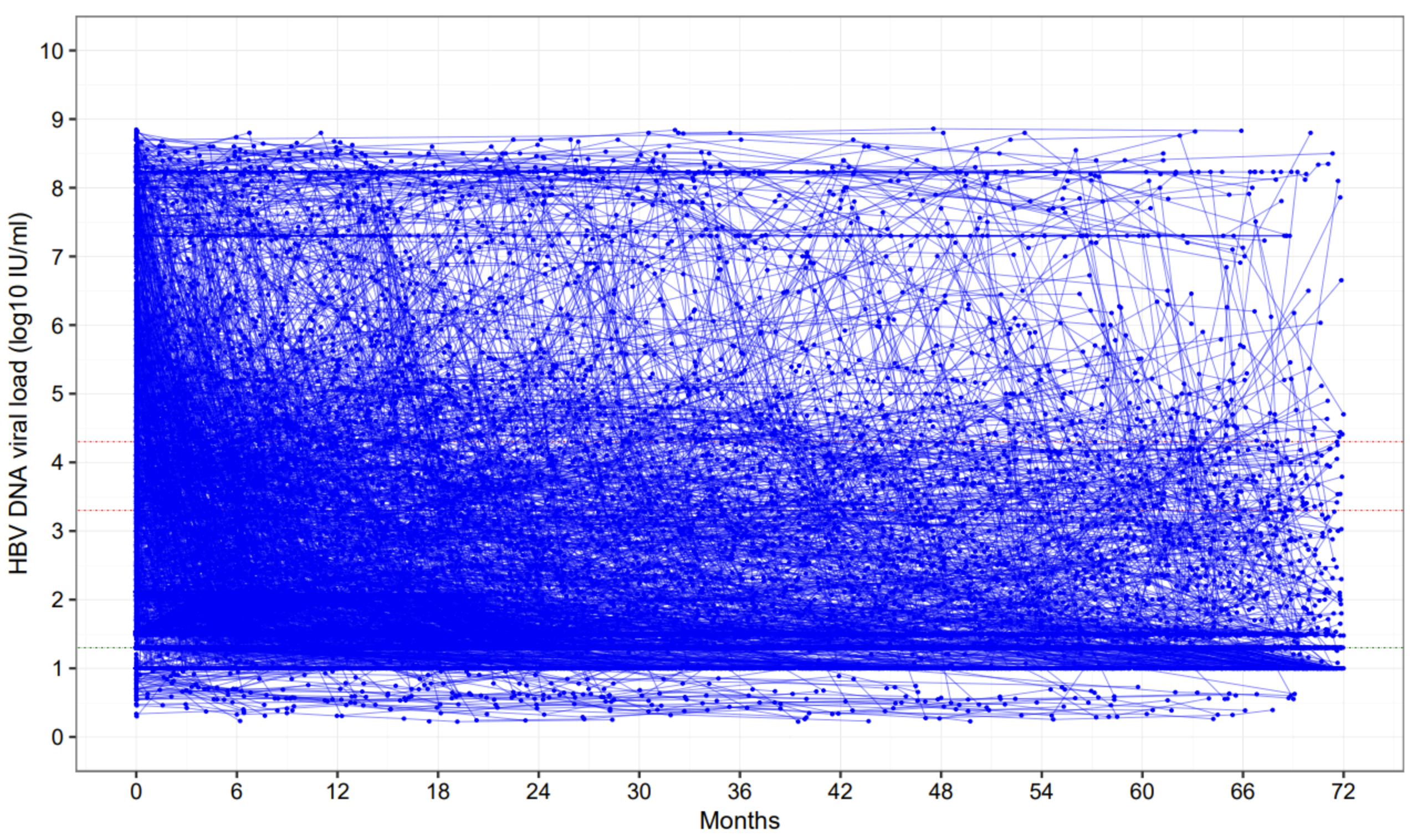

**Supplementary Figure S2.** Overview of individual virologic trajectories in 1885 adults with CHB on NA therapy (prior to the latent class analysis)

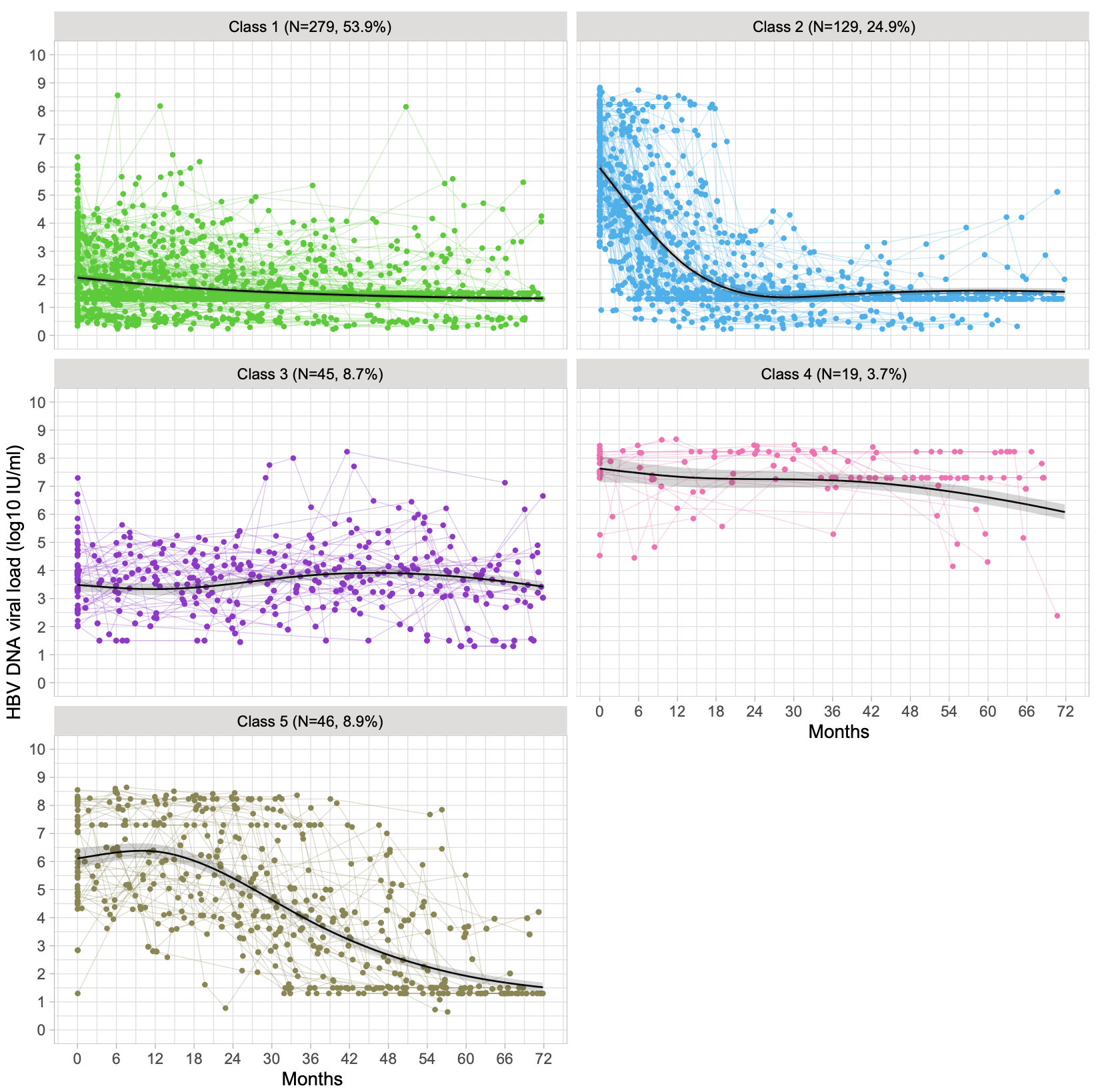

**Supplementary Figure S3.** Individual trajectories of HBV DNA viral load (VL) against the identified five VL trajectory patterns (‘classes 1-5’) for chronic hepatitis B patients on treatment in validation cohort (n=518). *Individual VL trajectories of validation cohort were classified using the estimated model based on derivation cohort. Dots represent the real values of VL, and solid lines with shading area represent the predicted VL trajectory patterns with 95% confidence intervals.*

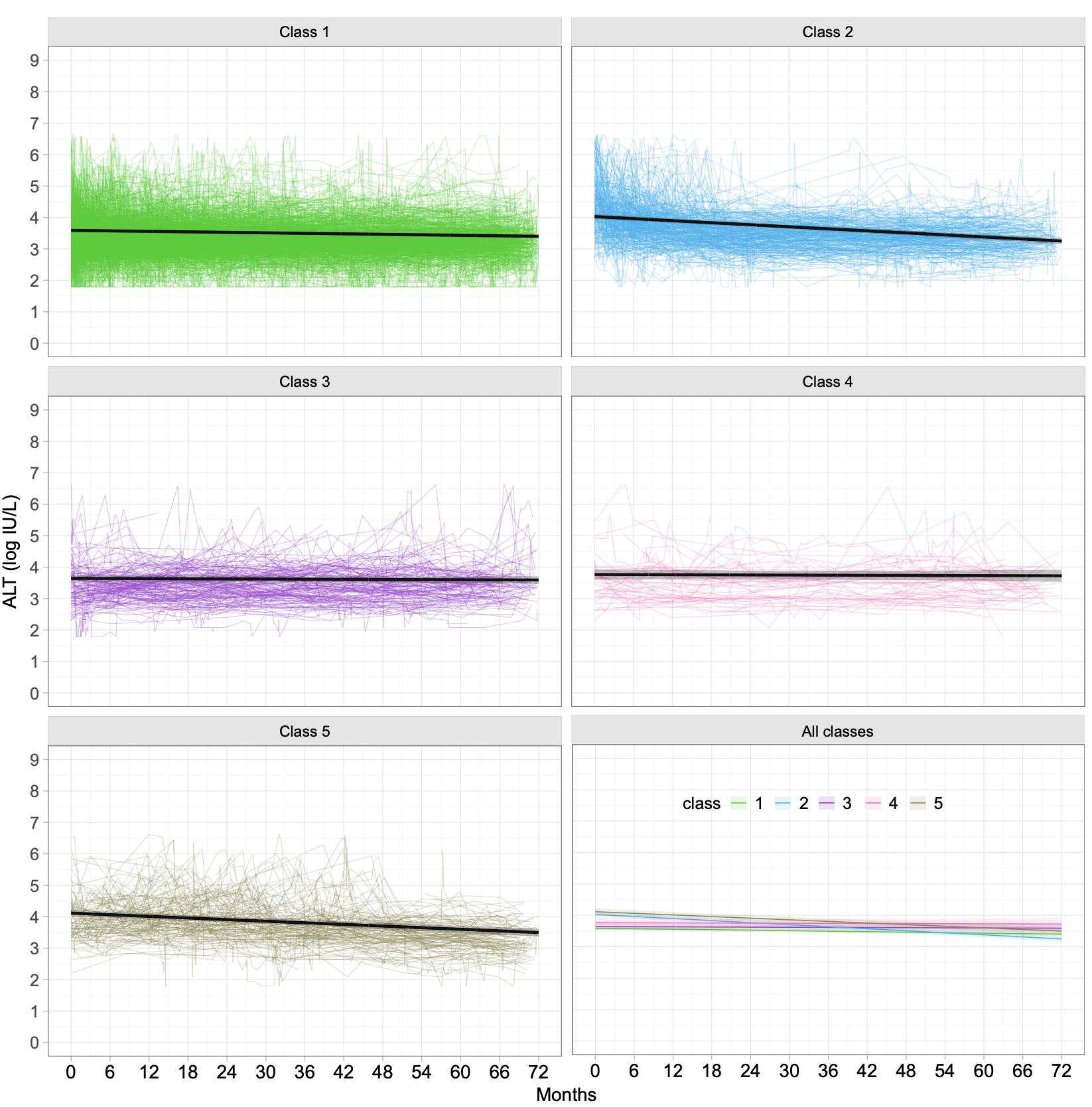

**Supplementary Figure S4**. Longitudinal trend of ALT levels (log IU/L) stratified by VL trajectory patterns in overall study cohort (n=1883, combining derivation and validation cohorts according to their identified VL trajectories, ALT data unavailable for three patients). *The trend was assessed by linear mixed effects models, considering the repeated measurements of each individual patient* and adjusted for age, sex, and ethnicity. *Coloured bands of the solid lines in last panel represent 95% confidence intervals.*

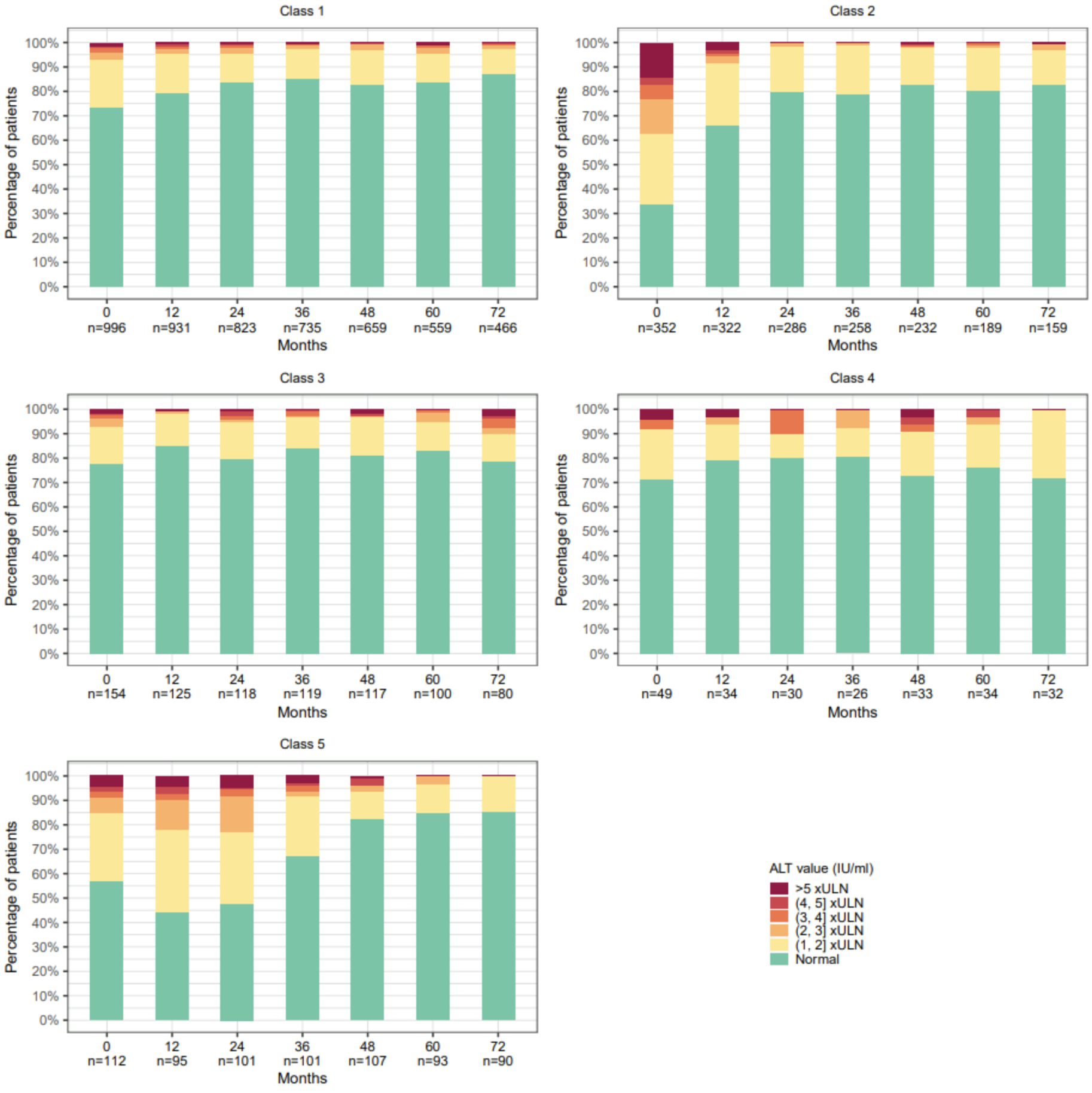

**Supplementary Figure S5.** Stratifications of ALT levels over time for patients with distinct virologic trajectory patterns. *ALT, Alanine aminotransferase. ULN, upper limit of normal.*

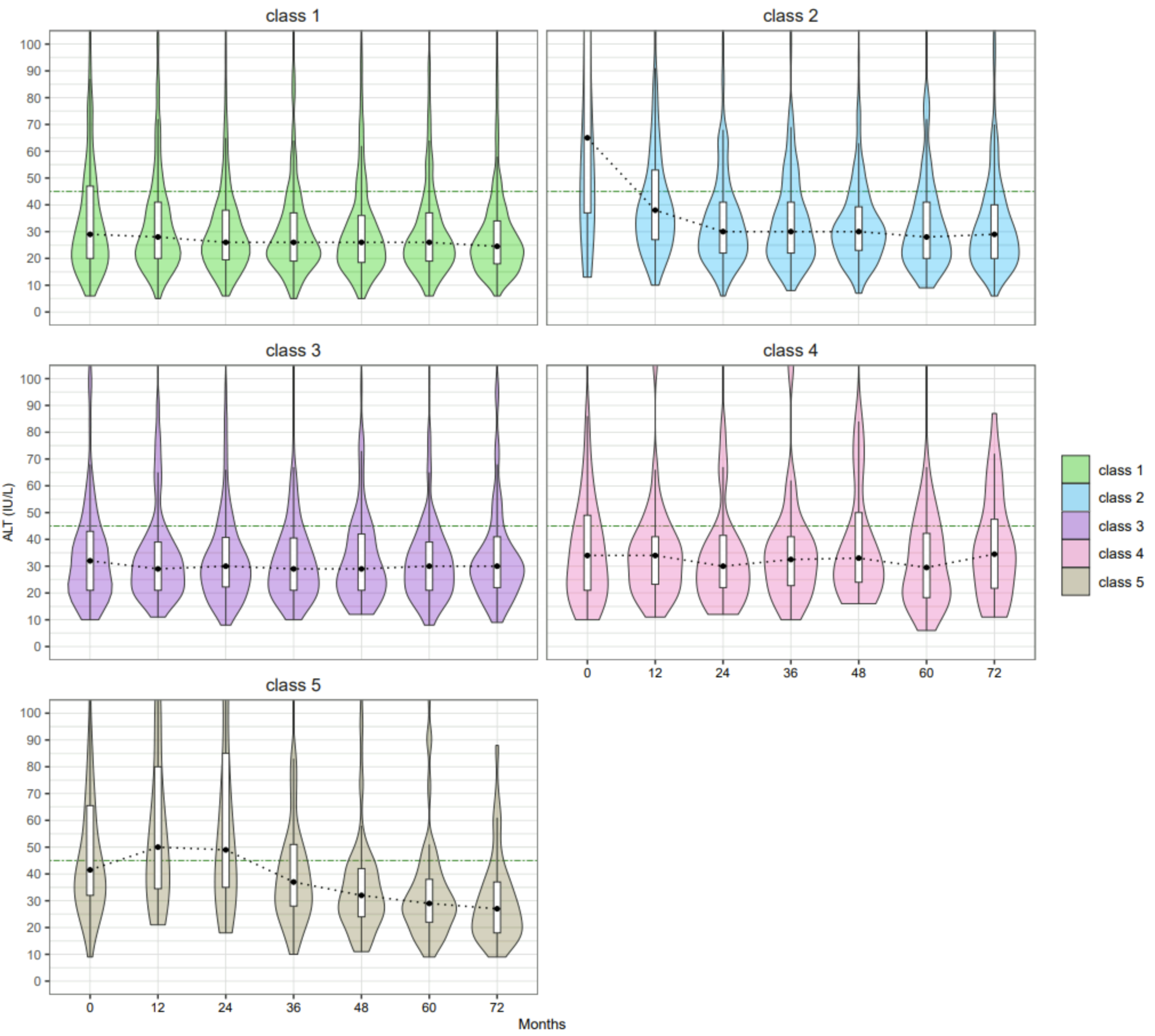

**Supplementary Figure S6**. Distribution of ALT levels at each time point for patients with distinct virologic trajectory patterns. *Green dash-dotted lines indicate the upper limit of normal (ULN) of ALT. Black dotted lines indicate the median values of ALT at each time point. The box represents the median and interquartile range, and the whiskers represent the adjacent values, which are within 1.5 times the interquartile range.*

**Supplementary Table S1**. Laboratory parameters used in this study

| Category | Laboratory parameter | Full name |
| --- | --- | --- |
| HBV lab tests | HBsAg | Hepatitis B surface antigen |
|  | HBV DNA VL | HBV DNA viral load |
|  | HBeAg | Hepatitis B e-Antigen |
|  | Anti-HBe | Anti-Hepatitis B e-antigen |
| Liver biochemistry and other tests reflecting liver health | ALT | Alanine aminotransferase |
|  | AST | Aspartate aminotransferase |
|  | Albumin |  |
|  | ALP | Alkaline phosphatase |
|  | Bilirubin |  |
|  | Platelets |  |
| Renal function | eGFR | Estimated Glomerular Filtration Rate |
|  | Urea |  |
| Virology tests for other chronic viral coinfections inference | HIV | Human immunodeficiency virus |
|  | HCV | Hepatitis C virus |
|  | HDV | Hepatitis D virus |

**Supplementary Table S2**. Calculation of discrimination and entropy

| **Metrics** | **Formula** | **Note** |
| --- | --- | --- |
| Discrimination | $\frac{\sum_{i=1}^{N} max\{P_{i}^{k}, k=1, 2, 3,K\}}{N}$ | $K$ represents the number of classes, $N$ indicates the number of patients included in the cohort for modelling. $P_{i}^{k}$ indicates the probability of patient $i$ being assigned to class $k$. |
| Entropy | $1 + \sum_{i=1}^{N} \frac{\sum_{k}^{K} log(P_{i}^{k})\cdot P_{i}^{k}}{N \cdot log(K)}$ |  |

**Supplementary Table S3**. Performance of the models with different number of classes of VL trajectories in the derivation cohort.

| Number of classes | BIC | SABIC | AIC | Entropy | Proportion for each class |
| --- | --- | --- | --- | --- | --- |
| 1 | 55654 | 55629 | 55612 | 1 | 100% |
| 2 | 54017 | 53954 | 53913 | 0.86 | 78.3%: 21.7% |
| 3 | 53212 | 53110 | 53045 | 0.88 | 72.7%: 19.6%: 7.7% |
| 4 | 52361 | 52221 | 52131 | 0.91 | 66.9%: 18.4%: 7.0%: 7.7% |
| 5 | 52124 | 51946 | 51832 | 0.90 | 60.5%: 18.6%: 10.2%: 3.2%: 7.5% |
| 6 | 51868 | 51652 | 51513 | 0.90 | 59.6%: 18.5%: 3.3%: 9.1%: 3.1%: 6.3% |

**Supplementary Table S4**. Classification performance of final estimated model in the derivation and validations cohorts

| **Assessment metrics** | **Derivation cohort** | **Validation cohort** |
| --- | --- | --- |
| Discrimination | 0.93 | 0.9287 |
| Entropy | 0.90 | 0.8815 |
| APPA for each class | 0.9509: 0.9221: 0.8543: 0.9436: 0.9330 | 0.9370: 0.9224: 0.8643: 0.9751: 0.9403 |
| Proportions per class | 60.5%: 18.6%: 10.2%: 3.2%: 7.5% | 53.9%: 24.9%: 8.7%: 3.7%: 8.9% |

APPA, average posterior probability assignment

**Supplementary Table S5.** Characteristics of patients at presentation stratified by the virologic trajectory patterns identified by latent class mixed modelling in the derivation cohort (n=1367)

| **Characteristics** | **Class 1**  **N=827 (60.5%)** | **Class 2**  **N=254 (18.6%)** | **Class 3**  **N=140 (10.2%)** | **Class 4**  **N=44 (3.2%)** | **Class 5**  **N=102 (7.5%)** | ***p* value** |
| --- | --- | --- | --- | --- | --- | --- |
|  | Long term suppression | Timely virological suppression | Persistent moderate viraemia | Persistent high-level viraemia | Slow virological suppression |  |
| Sex, male | 846 (61.9) | 66 (47.1) | 526 (63.6) | 179 (70.5) | 9 (20.5) | **<0.001** |
| Age, years | 47 [37, 58] | 44 [34, 55] | 38 [33, 45] | 30 [23, 35] | 36 [28, 46] | **<0.001** |
| Age group, years |  |  |  |  |  | **<0.001** |
| 18-24 | 16 (1.9) | 7 (2.8) | 4 (2.9) | 14 (31.8) | 14 (13.7) |  |
| 25-34 | 149 (18.0) | 62 (24.4) | 43 (30.7) | 18 (40.9) | 32 (31.4) |  |
| 35-44 | 187 (22.6) | 59 (23.2) | 57 (40.7) | 10 (22.7) | 25 (24.5) |  |
| 45-54 | 205 (24.8) | 59 (23.2) | 30 (21.4) | 2 (4.5) | 19 (18.6) |  |
| 55-64 | 147 (17.8) | 43 (16.9) | 5 (3.6) | 0 (0.0) | 11 (10.8) |  |
| 65-74 | 94 (11.4) | 17 (6.7) | 1 (0.7) | 0 (0.0) | 1 (1.0) |  |
| >=75 | 29 (3.5) | 7 (2.8) | 0 (0.0) | 0 (0.0) | 0 (0.0) |  |
| Ethnic group |  |  |  |  |  |  |
| Asian | 211 (30.8) | 86 (41.7) | 62 (47.7) | 23 (67.6) | 36 (43.4) | **<0.001** |
| Black | 175 (25.5) | 34 (16.5) | 19 (14.6) | 6 (17.6) | 9 (10.8) |  |
| White | 172 (25.1) | 44 (21.4) | 18 (13.8) | 1 (2.9) | 25 (30.1) |  |
| Mixed/other ethnicity | 127 (18.5) | 42 (20.4) | 31 (23.8) | 4 (11.8) | 13 (15.7) |  |
| Not reported | 142 (17.2) | 48 (18.9) | 10 (7.1) | 10 (22.7) | 19 (18.6) |  |
| IMD (decile) | 4.0 [2.0, 6.0] | 4.0 [2.0, 6.0] | 4.0 [2.0, 7.0] | 3.0 [1.0, 6.0] | 3.0 [1.8, 6.0] | **0.015** |
| IMD not reported | 66 (8.0) | 23 (9.1) | 52 (37.1) | 5 (11.4) | 10 (9.8) | **<0.001** |
| HBeAg status |  |  |  |  |  |  |
| Negative | 323 (80.8) | 98 (53.8) | 90 (89.1) | 3 (7.5) | 27 (32.5) | **<0.001** |
| Positive | 77 (19.2) | 84 (46.2) | 11 (10.9) | 37 (92.5) | 56 (67.5) |  |
| Not available | 427 (51.6) | 72 (28.3) | 39 (27.9) | 4 (9.1) | 19 (18.6) |  |
| Anti-HBe status |  |  |  |  |  |  |
| Negative | 125 (26.2) | 74 (41.1) | 15 (14.6) | 37 (92.5) | 54 (65.9) | **<0.001** |
| Positive | 353 (73.8) | 106 (58.9) | 88 (85.4) | 3 (7.5) | 28 (34.1) |  |
| Not available | 349 (42.2) | 74 (29.1) | 37 (26.4) | 4 (9.1) | 20 (19.6) |  |
| HBV VL, log10 IU/ml | 1.5 [1.3, 2.5] | 6.3 [5.1, 7.7] | 3.6 [2.8, 4.5] | 8.2 [7.5, 8.5] | 6.3 [4.4, 8.2] | **<0.001** |
| HBV VL category, IU/ml |  |  |  |  |  | **<0.001** |
| <20 | 118 (14.3) | 0 (0.0) | 1 (0.7) | 0 (0.0) | 1 (1.0) |  |
| 20 - <2000 | 590 (71.3) | 3 (1.2) | 52 (37.1) | 2 (4.5) | 6 (5.9) |  |
| 2000 - <20,000 | 77 (9.3) | 15 (5.9) | 50 (35.7) | 0 (0.0) | 13 (12.7) |  |
| >=20,000 | 42 (5.1) | 236 (92.9) | 37 (26.4) | 42 (95.5) | 82 (80.4) |  |
| ALT, IU/L | 28 [19, 45] | 70 [38, 147] | 31 [21, 40] | 35 [25, 50] | 42 [33, 73] | **<0.001** |
| AST, IU/L | 30 [24, 40] | 48 [34, 79] | 27 [23, 32] | 27 [22, 51] | 32 [28, 54] | **<0.001** |
| Platelets, 10^9/L | 199 [158, 248] | 193 [155, 231] | 218 [169, 257] | 203 [177, 247] | 221 [178, 270] | **0.001** |
| Albumin, g/L | 40.0 [36.0, 43.0] | 40.0 [37.0, 42.0] | 43.0 [39.2, 45.8] | 39.0 [35.8, 43.2] | 41.0 [38.0, 43.0] | **<0.001** |
| ALP, IU/L | 79.0 [62.0, 98.0] | 80.0 [61.0, 103.5] | 64.0 [52.0, 80.8] | 64.5 [60.0, 78.2] | 70.5 [61.0, 88.2] | **<0.001** |
| Bilirubin, µmol/L | 9.0 [6.0, 13.0] | 11.0 [7.5, 15.0] | 8.5 [6.0, 12.0] | 10.0 [6.0, 11.5] | 10.0 [7.5, 13.0] | **0.001** |
| eGFR category, mL/min/1.73 m^2^ |  |  |  |  |  | **<0.001** |
| >=90 | 251 (40.2) | 75 (43.1) | 53 (54.1) | 9 (75.0) | 37 (64.9) |  |
| >=60 & <=89 | 265 (42.5) | 88 (50.6) | 38 (38.8) | 3 (25.0) | 16 (28.1) |  |
| <=59 | 108 (17.3) | 11 (6.3) | 7 (7.1) | 0 (0.0) | 4 (7.0) |  |
| Not available | 203 (24.5) | 80 (31.5) | 42 (30.0) | 32 (72.7) | 45 (44.1) |  |
| Urea, mmol/L | 5.0 [4.0, 6.5] | 4.8 [3.9, 5.8] | 4.8 [3.9, 5.9] | 3.6 [3.2, 4.4] | 5.1 [4.1, 6.0] | **<0.001** |
| Urea, not available | 39 (4.7) | 16 (6.3) | 27 (19.3) | 12 (27.3) | 25 (24.5) |  |
| Treatment regimens |  |  |  |  |  | **<0.001** |
| TDF | 457 (56.2) | 186 (74.1) | 94 (67.6) | 33 (75.0) | 58 (60.4) |  |
| ETV | 161 (19.8) | 30 (12.0) | 24 (17.3) | 3 (6.8) | 11 (11.5) |  |
| ETV+TDF | 39 (4.8) | 19 (7.6) | 9 (6.5) | 3 (6.8) | 12 (12.5) |  |
| LAM/ADE+TDF | 29 (3.6) | 6 (2.4) | 2 (1.4) | 1 (2.3) | 6 (6.2) |  |
| LAM/ADE+ETV | 14 (1.7) | 0 (0.0) | 2 (1.4) | 0 (0.0) | 1 (1.0) |  |
| Other regimens | 113 (13.9) | 10 (4.0) | 8 (5.8) | 4 (9.1) | 8 (8.3) |  |

*Data are the number (%) or median [IQR]. IMD, Index of Multiple Deprivation; VL, viral load; ALT, alanine aminotransferase; TDF, tenofovir disoproxil fumarate; ETV, entecavir; LAM, lamivudine; ADE, adefovir. Comparison was conducted across non-missing categories for a categorical variable.*

**Supplementary Table S6.** Characteristics of patients at presentation stratified by the virologic trajectory patterns identified by latent class mixed modelling in the validation cohort (n=518).

| **Characteristics** | **Class 1**  **N =279 (53.9%)** | **Class 2**  **N=129 (24.9%)** | **Class 3**  **N=45 (8.7%)** | **Class 4**  **N=19 (3.7%)** | **Class 5**  **N=46 (8.9%)** |
| --- | --- | --- | --- | --- | --- |
|  | Long term suppression | Timely virological suppression | Persistent moderate viraemia | Persistent high-level viraemia | Slow virological suppression |
| Sex, male | 189 (67.7) | 87 (67.4) | 26 (57.8) | 11 (57.9) | 25 (54.3) |
| Age, years | 43 [35, 55] | 39 [32, 48] | 39 [33, 46] | 31 [24, 38] | 36 [31, 48] |
| Age group, years |  |  |  |  |  |
| 18-24 | 3 (1.1) | 9 (7.0) | 1 (2.2) | 5 (26.3) | 4 (8.7) |
| 25-34 | 64 (22.9) | 37 (28.7) | 16 (35.6) | 7 (36.8) | 14 (30.4) |
| 35-44 | 87 (31.2) | 39 (30.2) | 14 (31.1) | 4 (21.1) | 16 (34.8) |
| 45-54 | 54 (19.4) | 25 (19.4) | 9 (20.0) | 1 (5.3) | 5 (10.9) |
| 55-64 | 49 (17.6) | 15 (11.6) | 5 (11.1) | 2 (10.5) | 7 (15.2) |
| 65-74 | 17 (6.1) | 1 (0.8) | 0 (0.0) | 0 (0.0) | 0 (0.0) |
| >=75 | 5 (1.8) | 3 (2.3) | 0 (0.0) | 0 (0.0) | 0 (0.0) |
| Ethnic group |  |  |  |  |  |
| Asian | 107 (44.6) | 46 (44.2) | 22 (51.2) | 9 (56.2) | 22 (52.4) |
| Black | 44 (18.3) | 16 (15.4) | 6 (14.0) | 2 (12.5) | 3 (7.1) |
| White | 73 (30.4) | 33 (31.7) | 12 (27.9) | 3 (18.8) | 12 (28.6) |
| Mixed/other ethnicity | 16 (6.7) | 9 (8.7) | 3 (7.0) | 2 (12.5) | 5 (11.9) |
| Not reported | 39 (14.0) | 25 (19.4) | 2 (4.4) | 3 (15.8) | 4 (8.7) |
| IMD (decile) | 6.0 [4.0, 9.0] | 7.0 [5.0, 9.0] | 7.0 [5.0, 9.0] | 6.0 [5.0, 8.0] | 6.0 [4.0, 9.0] |
| IMD category |  |  |  |  |  |
| 20% most deprived | 28 (10.6) | 12 (9.8) | 2 (4.7) | 1 (5.6) | 5 (11.4) |
| 20% to 40% | 49 (18.6) | 18 (14.8) | 7 (16.3) | 3 (16.7) | 8 (18.2) |
| 40% to 60% | 57 (21.7) | 26 (21.3) | 12 (27.9) | 8 (44.4) | 11 (25.0) |
| 60% to 80% | 61 (23.2) | 31 (25.4) | 9 (20.9) | 2 (11.1) | 7 (15.9) |
| 20% least deprived | 68 (25.9) | 35 (28.7) | 13 (30.2) | 4 (22.2) | 13 (29.5) |
| Not reported | 16 (5.7) | 7 (5.4) | 2 (4.4) | 1 (5.3) | 2 (4.3) |
| HBeAg status |  |  |  |  |  |
| Negative | 169 (80.1) | 60 (48.8) | 26 (78.8) | 0 (0.0) | 18 (47.4) |
| Positive | 42 (19.9) | 63 (51.2) | 7 (21.2) | 17 (100.0) | 20 (52.6) |
| Not available | 68 (24.4) | 6 (4.7) | 12 (26.7) | 2 (10.5) | 8 (17.4) |
| Anti-HBe status |  |  |  |  |  |
| Negative | 51 (25.0) | 64 (52.0) | 6 (18.2) | 16 (94.1) | 18 (50.0) |
| Positive | 153 (75.0) | 59 (48.0) | 27 (81.8) | 1 (5.9) | 18 (50.0) |
| Not available | 75 (26.9) | 6 (4.7) | 12 (26.7) | 2 (10.5) | 10 (21.7) |
| HBV VL, log10 IU/ml | 2.1 [1.4, 3.4] | 6.4 [5.0, 7.7] | 3.9 [3.2, 4.6] | 8.0 [7.5, 8.2] | 7.1 [5.1, 8.1] |
| HBV VL category, IU/ml |  |  |  |  |  |
| <20 | 46 (16.5) | 0 (0.0) | 0 (0.0) | 0 (0.0) | 0 (0.0) |
| 20 - <2000 | 173 (62.0) | 1 (0.8) | 13 (28.9) | 0 (0.0) | 2 (4.3) |
| 2000 - <20,000 | 44 (15.8) | 14 (10.9) | 18 (40.0) | 0 (0.0) | 0 (0.0) |
| >=20,000 | 16 (5.7) | 114 (88.4) | 14 (31.1) | 19 (100.0) | 44 (95.7) |
| ALT, IU/L | 33.0 [23.0, 51.0] | 58.0 [34.0, 102.0] | 34.5 [22.0, 44.5] | 34.0 [17.5, 49.5] | 39.5 [30.2, 62.8] |
| AST, IU/L | 35.5 [27.0, 46.8] | 42.5 [34.5, 66.5] | 26.5 [20.5, 36.5] | 15.0 [15.0, 15.0] | 35.0 [30.0, 56.0] |
| Platelets, 10^9/L | 185.0 [150.0, 232.0] | 206.0 [168.0, 239.0] | 217.0 [174.0, 230.0] | 203.0 [193.0, 239.0] | 199.5 [179.8, 232.0] |
| Albumin, g/L | 40.0 [38.0, 43.0] | 40.0 [35.0, 42.0] | 41.0 [40.0, 44.0] | 36.0 [31.0, 40.0] | 39.0 [35.0, 41.2] |
| ALP, IU/L | 74.0 [61.0, 96.0] | 84.0 [65.0, 108.0] | 67.5 [59.5, 79.8] | 87.0 [71.0, 107.0] | 71.5 [59.5, 88.5] |
| Bilirubin, µmol/L | 11.0 [8.0, 14.0] | 11.0 [8.0, 15.0] | 10.0 [6.8, 16.5] | 7.0 [5.0, 9.0] | 9.5 [6.8, 12.2] |
| eGFR category, mL/min/1.73 m^2^ |  |  |  |  |  |
| >=90 | 146 (59.3) | 72 (64.9) | 13 (41.9) | 8 (80.0) | 15 (44.1) |
| >=60 & <=89 | 85 (34.6) | 37 (33.3) | 16 (51.6) | 2 (20.0) | 19 (55.9) |
| <=59 | 15 (6.1) | 2 (1.8) | 2 (6.5) | 0 (0.0) | 0 (0.0) |
| Not available | 33 (11.8) | 18 (14.0) | 14 (31.1) | 9 (47.4) | 12 (26.1) |
| Urea, mmol/L | 5.0 [4.2, 6.1] | 4.9 [4.0, 5.8] | 4.6 [3.9, 5.0] | 4.2 [3.9, 4.8] | 4.9 [4.2, 5.8] |
| Treatment regimens |  |  |  |  |  |
| TDF | 94 (33.7) | 64 (49.6) | 21 (46.7) | 7 (36.8) | 15 (32.6) |
| ETV | 73 (26.2) | 38 (29.5) | 15 (33.3) | 5 (26.3) | 17 (37.0) |
| ETV+TDF | 25 (9.0) | 19 (14.7) | 8 (17.8) | 6 (31.6) | 10 (21.7) |
| LAM/ADE+TDF | 28 (10.0) | 2 (1.6) | 1 (2.2) | 0 (0.0) | 0 (0.0) |
| LAM/ADE+ETV | 33 (11.8) | 1 (0.8) | 0 (0.0) | 0 (0.0) | 0 (0.0) |
| Other regimens | 26 (9.3) | 5 (3.9) | 0 (0.0) | 1 (5.3) | 4 (8.7) |

*Data are the number (%) or median [IQR]. IMD, Index of Multiple Deprivation; VL, viral load; ALT, alanine aminotransferase; TDF, tenofovir disoproxil fumarate; ETV, entecavir; LAM, lamivudine; ADE, adefovir. Significance tests were not conducted in the validation cohort due to small numbers in some classes.*

**Supplementary Table S7.** Comparisons of probability of being liver fibrosis and cirrhosis-free status over time for classes with distinct VL trajectory patterns

|  | Class 1 | Class 2 | Class 3 | Class 4 |
| --- | --- | --- | --- | --- |
| Class 2 | 0.727 | **-** | **-** | **-** |
| Class 3 | **0.003** | **0.009** | **-** |  |
| Class 4 | 0.340 | 0.397 | 0.466 | **-** |
| Class 5 | **0.003** | **0.034** | **<0.0001** | **0.029** |

*Data were p values. The pairwise comparisons were based on log-rank test.*

**Supplementary** **Table S8.** Univariate and multivariate Cox proportional-hazards models investigating associations of different VL trajectory patterns with liver disease progression to fibrosis and/or cirrhosis among adults with chronic hepatitis B on NAs therapy

|  | **Univariate analysis** | | **Multivariate analysis** | |
| --- | --- | --- | --- | --- |
| **Variables** | **Crude HR (95% CIs)** | ***p* value** | **Adjusted HR (95% CI)** | ***p* value** |
| VL trajectory |  |  |  |  |
| Class 1 (reference) | 1.0 |  | 1.0 |  |
| Class 2 | 1.05 (0.79 - 1.40) | 0.734 | 1.08 (0.77 - 1.51) | 0.654 |
| Class 3 | 0.53 (0.35 - 0.81) | **0.004** | 0.78 (0.50 - 1.23) | 0.286 |
| Class 4 | 0.75 (0.42 - 1.35) | 0.338 | 1.55 (0.79 - 3.01) | 0.199 |
| Class 5 | 1.63 (1.18 - 2.25) | **0.003** | 2.24 (1.55 - 3.24) | **<0.001** |
| Age group |  |  |  |  |
| 18-24 years | 1.54 (0.87 - 2.72) | 0.137 | 1.64 (0.9 - 3.01) | 0.109 |
| 25-34 years (reference) | 1.0 |  | 1.0 |  |
| 35-44 years | 1.12 (0.8 - 1.58) | 0.515 | 1.13 (0.79 - 1.6) | 0.499 |
| 45-54 years | 1.52 (1.08 - 2.14) | **0.016** | 1.31 (0.91 - 1.9) | 0.150 |
| 55-64 years | 2.34 (1.65 - 3.33) | **<0.001** | 1.95 (1.33 - 2.85) | **<0.001** |
| 65-74 years | 3.36 (2.16 - 5.23) | **<0.001** | 3.13 (1.93 - 5.07) | **<0.001** |
| >=75 years | 5.24 (2.74 - 10.02) | **<0.001** | 5.70 (2.84 - 11.44) | **<0.001** |
| Sex |  |  |  |  |
| Female (reference) | 1.0 |  | 1.0 |  |
| Male | 1.59 (1.25 - 2.02) | **<0.001** | 1.40 (1.06 - 1.84) | **0.016** |
| Ethnic group |  |  |  |  |
| White (reference) | 1.0 |  | 1.0 |  |
| Asian | 0.81 (0.61 - 1.09) | 0.163 | 0.91 (0.68 - 1.22) | 0.522 |
| Black | 1.17 (0.85 - 1.62) | 0.338 | 1.29 (0.92 - 1.82) | 0.141 |
| Mixed or other ethnicity | 1.02 (0.72 - 1.46) | 0.896 | 1.07 (0.74 - 1.55) | 0.732 |
| IMD |  |  |  |  |
| 20% most deprived | 1.01 (0.72 - 1.41) | 0.966 | 0.94 (0.67 - 1.31) | 0.719 |
| 20% to 40% | 1.14 (0.81 - 1.59) | 0.445 | 1.21 (0.86 - 1.70) | 0.275 |
| 40% to 60% (reference) | 1.0 |  | 1.0 |  |
| 60% to 80% | 1.21 (0.83 - 1.77) | 0.320 | 1.23 (0.86 - 1.78) | 0.262 |
| 20% least deprived | 1.23 (0.83 - 1.81) | 0.305 | 1.27 (0.86 - 1.90) | 0.231 |
| Coinfection^†^ (Yes) | 1.6 (1.01 - 2.51) | **0.043** | 1.09 (0.65 - 1.84) | 0.748 |
| HBeAg status (Positive) | 1.01 (0.77 - 1.32) | 0.939 | 1.01 (0.60 - 1.68) | 0.982 |
| Anti-HBe status (Positive) | 1.01 (0.79 - 1.29) | 0.928 | 1.52 (0.94 - 2.46) | 0.091 |
| Albumin, g/L | 0.94 (0.93, 0.96) | **<0.001** | 0.95 (0.93 - 0.97) | **<0.001** |
| ALP, 10 IU/L | 1.03 (1.02, 1.04) | **<0.001** | 1.03 (1.01 - 1.04) | **<0.001** |
| AST, 10 IU/L | 1.07 (1.05, 1.10) | **<0.001** | 1.07 (1.04 - 1.10) | **<0.001** |
| ALT, 10 IU/L | 1.01 (0.99, 1.01) | 0.199 | 0.99 (0.98 - 1.01) | 0.239 |
| Bilirubin, 10 µmol/L | 1.03 (0.99, 1.07) | 0.074 | 1.02 (0.97 - 1.08) | 0.422 |
| Platelets, 10*10^9/L | 0.94 (0.92, 0.96) | **<0.001** | 0.94 (0.92 - 0.96) | **<0.001** |
| Haemoglobin, 10 g/L | 1.01 (0.99, 1.03) | 0.113 | 1.01 (0.99 - 1.03) | 0.308 |
| HBV VL^‡^, log10 IU/ml | 0.98 (0.93, 1.03) | 0.40 |  |  |
| eGFR category |  |  |  |  |
| ≥90 mL/minute/1.73 m^2^ (reference) | 1.0 |  | 1.0 |  |
| ≥60 and ≤89 mL/minute/1.73 m^2^ | 1.09 (0.85 - 1.39) | 0.500 | 0.88 (0.68 - 1.13) | 0.303 |
| ≤59 mL/minute/1.73 m^2^ | 1.87 (1.36 - 2.58) | **<0.001** | 0.95 (0.60 - 1.51) | 0.834 |
| Urea, mmol/L | 1.06 (1.03 - 1.08) | **<0.001** | 1.00 (0.96 - 1.04) | 0.938 |
| Treatment regimens |  |  |  |  |
| TDF (reference) | 1.0 |  | 1.0 |  |
| ETV | 1.19 (0.89 - 1.58) | 0.239 | 0.92 (0.68 - 1.25) | 0.601 |
| ETV+TDF | 1.86 (1.32 - 2.62) | **<0.001** | 1.66 (1.16 - 2.37) | **0.006** |
| LAM/ADE+TDF | 1.62 (0.97 - 2.71) | 0.066 | 1.14 (0.65 - 1.98) | 0.655 |
| LAM/ADE+ETV | 2.21 (1.20 - 4.09) | **0.011** | 1.32 (0.69 - 2.55) | 0.404 |
| Other regimens | 1.56 (1.06 - 2.28) | **0.022** | 1.06 (0.69 - 1.62) | 0.791 |

*^†^ Coinfection with HIV, HCV, or HDV.*

*^‡^ The variable of HBV VL levels at baseline was not separately accounted for the multivariate analysis because the VL trajectory already incorporates the HBV VL levels at baseline. 1412 patients with data available for identifying liver fibrosis and cirrhosis were included for multivariate analysis. Class 1 (VL long term suppressed), Class 2 (persistent viraemia with moderate VL), Class 3 (VL suppressed as expected), Class 4 (VL non-suppressing with high VL), Class 5 (VL slowly suppressed). VL, viral load; ALT, Alanine transaminase; ALP, Alkaline phosphatase; HR, Hazards ratio, TDF, tenofovir disoproxil fumarate; ETV, entecavir; LAM, lamivudine; ADE, adefovir.*

**Supplementary Table S9.** Sensitivity analysis with only adjusting for age and sex for investigating associations of different VL trajectory patterns with liver disease progression to fibrosis and/or cirrhosis among adults with chronic hepatitis B on NAs therapy

| **Variables** | **Adjusted HR (95% CI)** | ***p* value** |
| --- | --- | --- |
| VL trajectory |  |  |
| Class 1 (reference) | 1.0 |  |
| Class 2 | 1.15 (0.86 – 1.53) | 0.354 |
| Class 3 | 0.72 (0.47 – 1.11) | 0.138 |
| Class 4 | 1.15 (0.62 – 2.13) | 0.665 |
| Class 5 | 1.93 (1.38 – 2.69) | **<0.001** |
| Age group |  |  |
| 18-24 years | 1.40 (0.78 – 2.51) | 0.264 |
| 25-34 years (reference) | 1.0 |  |
| 35-44 years | 1.10 (0.78 – 1.55) | 0.583 |
| 45-54 years | 1.50 (1.06 – 2.13) | **0.021** |
| 55-64 years | 2.20 (1.54 – 3.15) | **<0.001** |
| 65-74 years | 3.28 (2.09 – 5.15) | **<0.001** |
| >=75 years | 5.70 (2.95 – 11.01) | **<0.001** |
| Sex |  |  |
| Female (reference) | 1.0 |  |
| Male | 1.52 (1.19 – 1.94) | **<0.001** |

**Supplementary Table S10.** Predictive ability (AUC, Sensitivity, Specificity) of each single variable (VL trajectory, demographics, and biochemistry parameters) for liver fibrosis and cirrhosis

| **Variables** | **AUC** | **Sensitivity** | **Specificity** | **Accuracy** | **p-value**  **(compared to VL trajectory)** |
| --- | --- | --- | --- | --- | --- |
| VL trajectory | 0.658 (0.624-0.691) | 0.593 | 0.650 | 0.637 | - |
| Baseline age | 0.666 (0.632-0.699) | 0.581 | 0.655 | 0.637 | 0.62 |
| Sex | 0.635 (0.601-0.670) | 0.554 | 0.604 | 0.592 | 0.10 |
| Platelets | 0.678 (0.643-0.711) | 0.614 | 0.645 | 0.638 | 0.26 |
| AST | 0.642 (0.608-0.676) | 0.542 | 0.678 | 0.646 | 0.20 |
| Albumin | 0.655 (0.622-0.689) | 0.569 | 0.649 | 0.630 | 0.87 |
| ALP | 0.655 (0.621-0.689) | 0.566 | 0.676 | 0.650 | 0.83 |
| Treatment regimens | 0.653 (0.619-0.687) | 0.578 | 0.670 | 0.649 | 0.75 |

*Note: ALP, Alkaline phosphatase; ALT, Alanine transaminase; VL, viral load; ROC, receiver operating characteristic; AUC, the area under an ROC curve.*

**Supplementary Table S11.** Improvement of predictive ability of the addition of VL trajectories, other biochemistry parameters, and treatment regimens for liver fibrosis and cirrhosis

| **Predictors** | **AUC** | **Sensitivity** | **Specificity** | **Accuracy** | **p-value (compared to preceding row)** |
| --- | --- | --- | --- | --- | --- |
| Baseline age + sex | 0.669 (0.635, 0.703) | 0.590 | 0.636 | 0.625 | - |
| Baseline age + sex + PLT + AST + ALB + ALP | 0.731 (0.699, 0.763) | 0.639 | 0.688 | 0.676 | **<0.001** |
| Baseline age + sex + PLT + AST + ALB + ALP + Treatment regimens | 0.738 (0.706, 0.770) | 0.645 | 0.700 | 0.687 | 0.14 |
| Baseline age + sex + PLT + AST + ALB + ALP + Treatment regimens + VL trajectory | 0.756 (0.725, 0.787) | 0.684 | 0.714 | 0.707 | **<0.01** |

*All the models adjusted for other parameters that were included in the multivariate analysis, including ethnic group, IMD, HBeAg, anti-HBe, coinfection, ALT, Bilirubin, Hb, eGFR, and Urea.*

**Supplementary Table S12**. Classification performance in validation cohort using a smaller number of VL measurements with restrictions on minimum follow up duration (≥6 months)

|  | Use all VL measurements available | Use the first two VL measurements | Use the first three VL measurements | Use the first four VL measurements | Use the first five VL measurements |
| --- | --- | --- | --- | --- | --- |
| Number of patients | 518 | 260 | 378 | 394 | 359 |
| Follow up duration (months) of VL measurements, median [IQR] | 49 [25, 77] | 12 [7, 19] | 14 [9, 24] | 19 [12, 31] | 25 [17, 37] |
| Discrimination | 0.9287 | 0.8323 | 0.8268 | 0.8487 | 0.8667 |
| Entropy | 0.8815 | 0.7307 | 0.7350 | 0.7678 | 0.7922 |
| APPA for each class  (class 1: class 2: class 3: class 4: class 5) | 0.9370:  0.8643:  0.9224:  0.9751:  0.9403 | 0.8809:  0.6221:  0.8342:  0.8162:  0.7402 | 0.8845:  0.6523:  0.8365:  0.7958:  0.7594 | 0.8916  0.7016  0.8709  0.8729  0.7848 | 0.8865:  0.7657:  0.8936:  0.8841:  0.8283 |
| Proportions per class  (class 1: class 2: class 3: class 4: class 5) | 53.86%:  8.69%:  24.9%:  3.67%:  8.88% | 61.54%:  8.85%:  10.77%:  7.69%:  11.15% | 51.59%:  12.7%:  17.46%:  8.2%:  10.05% | 45.43%:  13.45%:  22.84%:  7.87%:  10.41% | 43.18%:  13.37%:  24.51%:  10.03%:  8.91% |

APPA, average of maximum posterior probability of assignments.

**Supplementary Table S13**. Classification performance in validation cohort using a smaller number of VL measurements without restriction on minimum follow up duration

|  | Use all VL measurements available | Use the first two VL measurements | Use the first three VL measurements | Use the first four VL measurements | Use the first six VL measurements |
| --- | --- | --- | --- | --- | --- |
| Number of patients | 518 | 518 | 469 | 421 | 369 |
| Follow up duration (months) of VL measurements, median [IQR] | 49 [25, 77] | 6 [3, 12] | 12 [6, 21] | 19 [11, 30] | 24 [16, 37] |
| Discrimination | 0.9287 | 0.7835 | 0.8160 | 0.8481 | 0.8666 |
| Entropy | 0.8815 | 0.6660 | 0.7160 | 0.7642 | 0.7909 |
| APPA for each class | 0.9370:  0.8643:  0.9224:  0.9751:  0.9403 | 0.861:  0.572:  0.719:  0.745:  0.661 | 0.8765: 0.6382: 0.8111: 0.7850:  0.7446 | 0.8897:  0.7022:  0.8689:  0.8734:  0.7786 | 0.8850:  0.7657  0.8933  0.8841  0.8283 |
| Proportions per class | 53.86%:  8.69%:  24.9%:  3.67%:  8.88% | 55.98%: 7.34%: 19.11%: 7.34%: 10.23% | 49.89%: 11.3%: 21.75%: 7.68%:  9.38% | 45.61%  12.83%  23.75%  7.6%  10.21% | 43.09%  13.01%  25.47%  9.76%  8.67% |

APPA, average of maximum posterior probability of assignments.

**Table S14**. Relationship between VL classes and EASL definition of virological breakthrough

|  | Virological breakthrough^†^ |
| --- | --- |
| Overall cohort (n=1885) | 120/1885 (6.4%) |
| Class 1 (n=1106) | 44/1106 (4.0%) |
| Class 2 (n=383) | 11/383 (2.9%) |
| Class 3 (n=185) | 32/185 (17.3%) |
| Class 4 (n=63) | 19/63 (30.2%) |
| Class 5 (n=148) | 14/148 (9.5%) |

^†^ *Virological breakthrough: a confirmed increase in HBV DNA level of more than 1 log10 IU/ml compared to the nadir HBV DNA level on-therapy.*
